# Artificial Intelligence in Biomedical Data Analysis: A Comparative Assessment of Large Language Models for Automated Clinical Trial Interpretation and Statistical Evaluation

**DOI:** 10.1101/2025.02.05.25321607

**Authors:** Ravikumar Komandur, Jon McDunn, Nikita Nair, Babacar Fall, Adam P. Dicker, Sean Khozin

**Affiliations:** Project Data Sphere, Cary, NC, 27513; CEO RoundTable on Cancer, Cary, NC, 27513; Sidney Kimmel Medical College at Thomas Jefferson University, Philadelphia, PA

**Keywords:** large language models, clinical trials, small cell lung cancer, ChatGPT, Gemini Advanced, chain-of-thought prompting, data analysis

## Abstract

**Background:** Clinical trials provide evidence of the efficacy and safety of experimental treatment regimens. Analysis of data from these trials is a time-intensive process traditionally requiring advanced multidisciplinary expertise in biomedicine, clinical research, biostatistics and data science. Large language models (LLMs), such as OpenAI’s GPT-4 and Google’s Gemini Advanced, present new opportunities for data analysis in medical research by leveraging natural language understanding and data interpretation capabilities.

**Objective:** This study investigates the ability of LLMs to analyze and report clinical trial results, starting with de-identified individual patient data. Here, we evaluate two LLMs for their ability to recapitulate the analysis of a clinical trial that evaluated LY2510924 in combination with carboplatin and etoposide for the treatment of extensive-stage small cell lung cancer (ES-SCLC). The main objectives are to (i) assess whether LLMs can be effectively used without specialized machine learning training and (ii) compare LLM-driven analyses to those conducted by experienced data scientists.

**Methods:** Data from the Project Data Sphere (PDS) platform were used, and multiple investigators employed both ChatGPT and Gemini Advanced for analysis. A chain-of-thought (CoT) prompting framework was applied to guide the LLMs through a systematic evaluation of baseline characteristics, progression-free survival (PFS), overall survival (OS), safety data, and biomarker information. Results were compared across investigators and LLMs to assess consistency.

**Results:** While LLMs could process the trial data and generate relevant insights, discrepancies were observed across the investigators’ analyses, particularly in primary and secondary endpoints. One investigator found a significant improvement in PFS with LY2510924, contradicting other results. Variations in reporting objective response rate (ORR) and adverse event analyses also highlighted challenges in reporting between different LLMs. These discrepancies may be due to differences in LLM capabilities and behaviors, prompting strategies, and potential model drift over time.

**Conclusion:** LLMs such as ChatGPT-4 and Gemini Advanced offer promising capabilities in clinical data analysis, though variability in results underscores the need for tailored CoT frameworks and specialized prompting strategies. Addressing issues such as model drift and ensuring consistent model versions are crucial for reliable application. LLMs also show potential to accelerate clinical research by drafting clinical trial reports, but further refinements are needed to ensure accuracy and consistency in their application. The observed discrepancies across LLM results and in comparison, to the expert-authored trial report highlight the need for highly trained subject matter experts to review and revise LLM-generated clinical trial analyses.

## 1.0 INTRODUCTION

Analyzing clinical trial data is crucial for assessing the efficacy and safety of new treatments. Traditionally, this process has required specialized expertise in biomedicine, clinical research, biostatistics and data science, often making it labor-intensive, time-consuming, and costly [1]. For clinicians and researchers lacking advanced training in data analytics, the complex statistical requirements can become significant barriers, leading to delays in translating research findings into clinical practice. Moreover, the increasing complexity of modern clinical trials, characterized by large datasets and multiple endpoints, exacerbates these challenges [2]. The integration of diverse primary and secondary data sources, which clinical trials increasingly rely upon, further underscores the need for advanced analytical tools to handle complex, heterogeneous data. Interventional clinical trials rely on meticulous record-keeping under exacting protocols, involving experts across multiple disciplines, including—disease biology, specialty clinical care, toxicology, translational sciences, biostatistics, bioanalytical sciences, regulatory affairs, and biomedical ethics. Each field contributes essential elements to trial design, ensuring that every aspect of a trial aligns with regulatory standards and scientific rigor to produce evidence on therapy efficacy and safety.

Advancements in artificial intelligence (AI), particularly large language models (LLMs), have shown promise in addressing these obstacles by streamlining data integration and analysis processes [2]. The adoption of frameworks like C-DISC [3] has standardized data collection and reporting, allowing structured interpretation across trials. Building upon these frameworks, AI-driven models like LLMs offer the potential for a prespecified evaluation and reporting system that could seamlessly integrate with existing trial structures. The advent of large language models (LLMs), such as OpenAI’s GPT-4 [4] and Google’s Gemini Advanced [5], has opened new avenues for data analysis in medical research [6]. LLMs have demonstrated capabilities in natural language understanding, data interpretation [7], and generating human-like text with reasoning capabilities. These abilities make them valuable tools for various applications, including summarizing medical literature and assisting in clinical decision-making [8, 9]. They have been successfully utilized across diverse fields, from basic science to clinical practice, for summarizing research papers, generating clinical documentation, and enhancing scientific writing accuracy and efficiency. Notably, LLMs have been explored for their potential to interpret complex results and identify patterns within datasets, which could revolutionize data analysis in clinical research [10].

Recent studies have demonstrated the potential of LLMs, such as ChatGPT Advanced Data Analysis (ADA) [11], to autonomously develop and implement machine learning models for clinical outcome predictions across various medical specialties. However, despite these advancements, there remains a gap in understanding how effectively LLMs can analyze raw clinical trial data without human intervention and whether they can produce results consistent with traditional statistical analyses [12]. In this study, we investigated the capacity of current LLMs to analyze data from a retrospective clinical trial evaluating the efficacy of LY2510924 combined with carboplatin and etoposide for the treatment of extensive-stage small-cell lung cancer (ES-SCLC) [913]. Our primary objective is twofold: first, to assess whether LLMs can be used intuitively without specialized machine learning training, and second, to determine whether they can produce results comparable to those achieved by experienced data scientists based on predefined criteria.

To evaluate the adequacy of the LLMs’ performance, we established specific criteria:

1. **Accuracy of Statistical Analyses:** The LLMs should perform appropriate statistical tests and report accurate results.
2. **Consistency with Original Trial Results:** The LLM-generated analyses should broadly align with the findings reported in the original clinical trial publication.
3. **Interpretation of Key Clinical Endpoints:** The LLMs should reasonably interpret and report on primary and secondary endpoints, including PFS, OS, ORR, and safety data.
4. **Reproducibility Across Investigators and Models:** The results should be consistent at a very high level when different investigators use the same LLMs and prompting frameworks.

By employing these criteria, we aim to determine the extent to which LLMs can replicate expert-level analyses of clinical trial data.

We hypothesized that large language models (LLMs) when guided by a structured chain-of-thought (CoT) prompting framework, can produce high-quality clinical trial reports by performing accurate and consistent analyses of clinical trial data. Leveraging the structure and quality of clinical trial datasets and the standard safety and efficacy assessments typically conducted, we anticipated that LLMs would replicate key findings, including baseline characteristics, efficacy endpoints, safety profiles, and biomarker analyses, aligning closely with traditional statistical analyses performed by experienced data scientists, even without specialized machine learning training.

We employed LLMs to draft an analytical code for generating statistical results and data visualizations commonly seen in clinical trials to achieve these objectives. We then compared these outputs with the published results of the trial. We aimed to demonstrate that advanced LLMs such as ChatGPT and Gemini Advanced can simplify clinical trial data analysis, potentially broadening their application in medical research.

Furthermore, we highlight the role of prompt engineering—in particular, chain-of-thought (CoT) prompting—in guiding LLMs to produce structured summaries and generate statistical outputs that accurately reflect significant components of clinical trial documentation. We employ a structured chain-of-thought (CoT) prompting framework to standardize the analytical approach and assess its effectiveness in guiding LLMs through complex data interpretation tasks [14]. By exploring the utility of LLMs in clinical trial data analysis, we hope to provide insights into their potential to accelerate research, reduce barriers to data interpretation, and ultimately contribute to improved patient outcomes. Our findings highlight the potential of LLMs to reproduce structured clinical trial documentation and present an efficient, scalable alternative for clinical trial reporting. These findings could have significant implications for the future integration of AI in clinical research, potentially making complex data analysis powered by LLMs accessible to non-experts, thus democratizing data analysis and enabling more rapid advancements in medical science [15].

## 2.0 MATERIALS AND METHODS

### 2.1 Dataset and Study Description

The datasets for this analysis were retrieved from the Project Data Sphere (PDS) Platform [16,17] and had been de-identified before storage to ensure patient privacy. Compared to the dataset used in the original study analysis [13], the PDS version underwent additional modifications to prevent a complete replication of the initial findings. Specifically, certain data points were either imputed or removed. Imputation involves replacing missing values with statistically plausible estimates derived from the available data. While this method can enhance dataset completeness, it can also introduce an element of uncertainty, particularly if the original missing values were non-random or correlated with specific patient subgroups. Additionally, the removal of key variables during dataset preparation may limit access to potentially influential data points.

These modifications may introduce variability in our re-analysis using LLMs. The absence of certain original variables or the presence of imputed values could lead to differences in statistical outcomes and interpretations compared to the original study. For example, missing data related to patient demographics, adverse events, or treatment response could impact the precision of our survival analysis and the assessment of progression-free survival (PFS) or overall survival (OS) outcomes independent of LLM performance.

The primary objective of the original study was to evaluate the efficacy of LY2510924 in combination with carboplatin and etoposide compared to carboplatin and etoposide alone in participants with extensive-stage Small Cell Lung Cancer (SCLC). Efficacy was primarily assessed by Progression-Free Survival (PFS) as per RECIST 1.1 criteria. Secondary objectives included the evaluation of overall survival (OS), objective response rate (ORR), duration of response (DOR), and safety. A total of 90 participants were randomized in a 1:1 ratio into one of two treatment arms:

- **Arm A:** LY2510924 + carboplatin + etoposide
- **Arm B:** Carboplatin + etoposide

### 2.2 Methods

We downloaded the clinical trial dataset from the PDS platform, which is available in SAS formats. The SAS files were converted into CSV format to make them compatible with prompting ChatGPT and Gemini Advanced. We used the ChatGPT-4 version and Gemini Advanced for our data analysis. We also utilized Jupyter Notebook with Python (v3.9) to generate survival curves, employing open-source libraries such as NumPy, SciPy, scikit-learn, and pandas to generate the OS and PFS curves.

A total of four investigators participated in the analysis: two from the PDS team and two from the Jefferson team. One investigator from each team used ChatGPT (Investigator 1 from PDS and Investigator 4 from Jefferson). In contrast, the other investigators from each team used Gemini Advanced (Investigator 2 from PDS and Investigator 3 from Jefferson). All investigators conducted the analysis independently with minimal consultation to ensure that the results reflected independent interpretations.

We adopted a chain-of-thought prompting template (as outlined in Supplementary Materials 1.1) for the analysis, with minor adjustments to the prompts guided by model feedback.

We uploaded background information into the LLMs as structured inputs before issuing prompts to elicit analytical outputs, effectively utilizing the models for data analysis. The background information was provided through (i) the study protocol detailing the clinical trial methodology, (ii) a data dictionary defining the dataset structure, variable meanings, data types, and permissible values, and (iii) various data files containing survival data, safety profiles, biomarkers, and other relevant clinical information. We did not apply any pre-training or fine-tuning to adapt the models to these specific datasets. Instead, our objective was to leverage their native capabilities, using the Chain-of-Thought (CoT) framework to structure their reasoning and analysis. Figure 1 provides an overview of the study design, including the data provisioning and analysis workflow.

**Figure 1.**
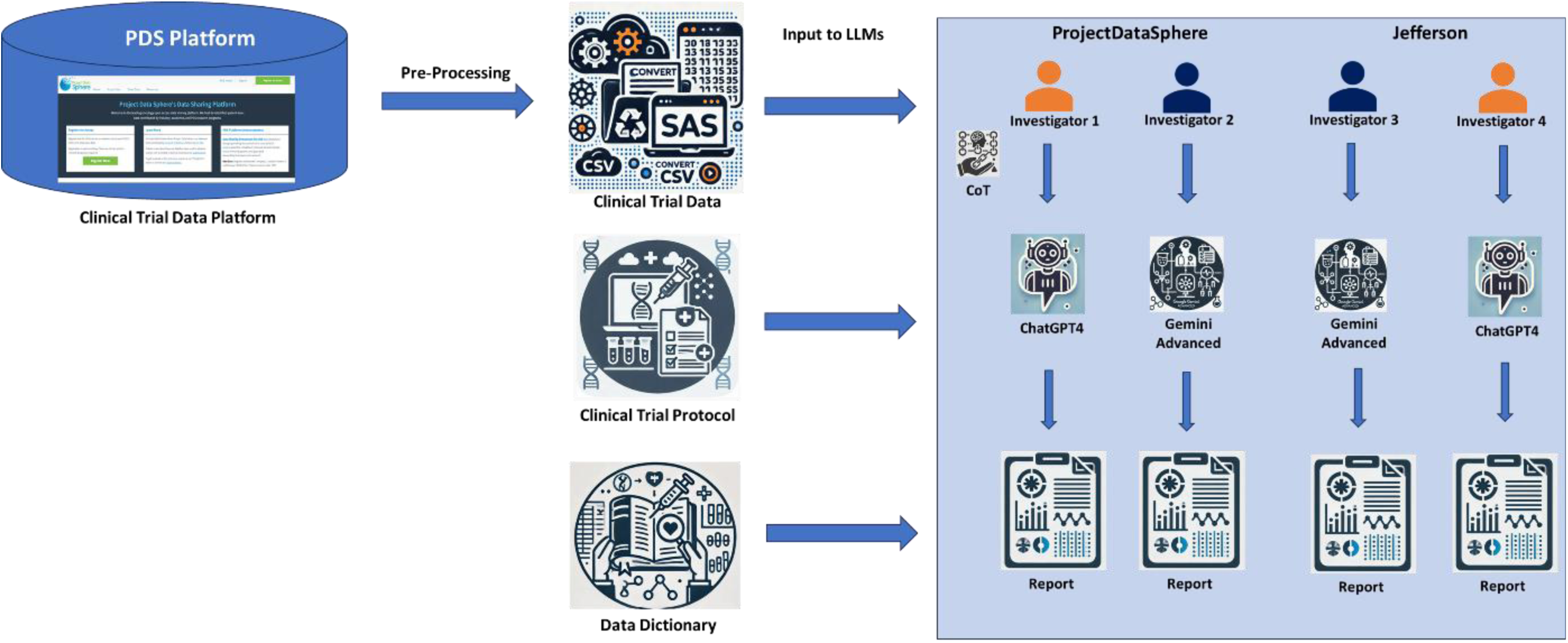
Schematic overview of the study workflow. Illustrating the process of data acquisition from the Project Data Sphere (PDS) platform, preparation and conversion of data files, assignment of investigators and Large Language Models (LLMs), utilization of the Chain-of-Thought (CoT) prompting framework for standardized analysis, independent data analysis by investigators using ChatGPT-4 and Gemini Advanced, generation of survival curves using Python libraries, compilation of results for comparison, and the derivation of conclusions and insights on the utility of LLMs in clinical trial data analysis.

### 2.3 Evaluation Criteria

To evaluate the performance of the LLMs in analyzing the clinical trial data, we established a set of predefined assessment criteria:

1. **Accuracy of Statistical Analyses:**

a. The LLMs should correctly select and perform appropriate statistical tests (e.g., Kaplan-Meier survival analysis).
2. **Consistency with Original Trial Results:**

a. The findings generated by the LLMs should be broadly consistent with those reported in the original clinical trial publication, particularly regarding primary and secondary endpoints.
3. **Correct Interpretation of Clinical Data:**

a. The LLMs should correctly interpret baseline characteristics, efficacy outcomes, and safety profiles.
b. Identify and report any significant differences or lack thereof between treatment arms.
4. **Reproducibility Across Investigators and Models:**

a. Analyses should yield consistent results to a reasonable extent, at least at a very high level, when performed by different investigators using the same LLMs and CoT prompting framework.
b. Any observed discrepancies should be attributable to minor variations in prompting or data interpretation rather than fundamental inconsistencies in model performance.
5. **Ability to Generate Appropriate Data Visualizations:**

a. When tasked, the LLMs should assist in generating accurate and relevant data visualizations (e.g., Kaplan-Meier curves).
6. **Identification of Limitations and Potential Biases:**

a. The LLMs should recognize limitations in the data or analysis, such as missing data, potential biases, or confounding factors.

These criteria provided a structured framework for evaluating the adequacy of the LLMs’ performance in replicating expert analyses of clinical trial data.

## 3.0 RESULTS

### 3.1 Baseline Characteristics Analysis

Four independent investigators independently analyzed the baseline characteristics of the clinical trial data that investigated the efficacy and safety of LY2510924 plus carboplatin/etoposide compared to carboplatin/etoposide alone in extensive-stage small cell lung cancer (ES-SCLC) using different Large Language Models (LLMs) to confirm that the treatment groups were comparable at the outset. This step was crucial to ensure that any observed differences in treatment outcomes were due to the treatment itself and not simply reflections of pre-existing differences between the groups. Their analyses focused on key demographic factors like age, sex, race, and ECOG performance status (a coarse-grained measure of patient health), providing a comprehensive picture of the patient population and laying the groundwork for a robust interpretation of the trial’s results.

The original publication of the clinical trial data reported that both treatment arms were similar regarding baseline demographic and clinical characteristics. However, the control arm (carboplatin/etoposide alone) had proportionally more females. Tumor CXCR4 expression at baseline was assessed via immunohistochemistry (IHC), with mean and median H-scores found to be comparable between arms.

Baseline analyses performed using LLMs showed broad agreement that the treatment groups were balanced for key demographic factors such as age and sex. However, there were inconsistencies in the results generated by the LLMs, despite all investigators working with the same input dataset. For example, ECOG performance status results were included in some outputs but were entirely absent in others. Similarly, LDH levels were reflected in only one set of results, and race was inconsistently addressed, with some outputs omitting it entirely. These variations likely stem from differences in how the LLMs processed and interpreted the input data, as guided by the investigators’ prompting strategies, rather than differences in the dataset itself.

Despite these inconsistencies, the overall LLM results consistently indicated that the randomization process successfully created balanced treatment groups. This reinforces confidence that any observed differences in treatment outcomes are likely attributable to the intervention rather than pre-existing imbalances.

*Table 1 summarizes the baseline characteristics analyzed by four independent investigators in a clinical trial comparing LY2510924 plus carboplatin/etoposide with carboplatin/etoposide alone for extensive-stage small cell lung cancer (ES-SCLC)*.

**Table 1.** Comparison of Baseline Characteristics Analyses by Four Independent Investigators.

| **Characteristic** | **Investigator 1**<br>**(ChatGPT-4)** | **Investigator 2**<br>**(Gemini Advanced)** | **Investigator 3**<br>**(Gemini Advanced)** | **Investigator 4**<br>**(ChatGPT-4)** |
| --- | --- | --- | --- | --- |
| **Age** | Mean age comparable<br>between groups<br>(p=0.60) | Missing data<br>prevented calculation | Mean age comparable<br>(p=0.88) | Groups comparable |
| **Sex** | No significant difference<br>(p=0.78) | No significant difference<br>(p=0.6987) | No significant difference<br>(p=0.65) | Not reported |
| **Race** | Not reported | No significant difference<br>(p=0.5477) | Slight imbalance, not<br>statistically significant<br>(p=0.13) | Not reported |
| **ECOG**<br>**Performance Status** | No significant difference<br>(p=0.87) | No significant difference<br>(p=0.7641) | Not reported | Data missing |
| **LDH Levels** | No significant difference<br>(p=0.82) | Not reported | Not reported | Not reported |

### 3.2 Analysis of Primary and Secondary Endpoints

To rigorously assess the efficacy of LY2510924 in combination with carboplatin/etoposide for the treatment of extensive-stage small cell lung cancer (ES-SCLC), we conducted analyses of primary and secondary endpoints from the clinical trial data. These analyses primarily focused on progression-free survival (PFS) and overall survival (OS), along with objective response rate (ORR) and duration of response (DOR).

#### 3.2.1 Original Publication Findings

The original publication of this clinical trial reported that adding LY2510924 to carboplatin/etoposide did not significantly improve PFS or OS. The ORR was numerically higher in the carboplatin/etoposide alone arm (81% vs. 74.5% with LY2510924), though this difference was not statistically significant.

#### 3.2.2 Investigator Analyses

Four independent investigators conducted separate analyses of the clinical trial data using LLMs. One investigator found a statistically significant improvement in PFS with the addition of LY2510924. In contrast, the other three investigators confirmed the original publication’s finding of no significant difference in PFS or OS.

Discrepancies were also observed in the ORR findings. One investigator reported a higher ORR in the LY2510924 combination arm, although not statistically significant. Another investigator reported an ORR of 0% in both arms, significantly deviating from the original publication and other investigators’ findings. This substantial deviation likely indicates an error in data interpretation or processing by the LLM, possibly due to ambiguity in the data or limitations in the model’s understanding of the specific context. Model drift could also influence these discrepancies, where updates or changes in the LLMs over time result in different outputs, even when using the same inputs.

#### 3.2.3 Subgroup Analyses

Subgroup analyses explored potential treatment effects within specific patient populations (based on ECOG performance status, LDH levels, age, and gender). However, these analyses did not reveal consistent or statistically significant differences between the treatment arms and subgroups.

#### 3.2.4 High-Point Summary

- **Inconclusive Results:** The analyses of primary and secondary endpoints yielded inconsistent results regarding the efficacy of LY2510924 in ES-SCLC.
- **PFS Discrepancy:** One investigator found a significant improvement in PFS, contradicting the original publication and the other investigators’ analyses.
- **ORR Variations:** Significant variations in ORR were observed across the different analyses.
- **No Subgroup Effects:** No consistent treatment effects were found within any of the examined subgroups.

*Table 2 summarizes the findings of the original publication and the four investigators’ analysis of primary and secondary endpoints*.

**Table 2.** Comparison of Primary and Secondary Endpoint Analyses.

| **Study** | **PFS** | **OS** | **ORR** | **DOR** | **Safety** | **Subgroup Analysis** |
| --- | --- | --- | --- | --- | --- | --- |
| **Original Publication** | No significant difference<br>(5.88 vs. 5.85 months) | No significant difference<br>(9.72 vs. 11.14 months) | 74.5% vs. 81% | No significant difference<br>(4.83 vs. 4.67 months) | Similar between arms,<br>but some AEs more<br>frequent in Arm A | Not reported |
| **Investigator 1** | Significant difference<br>(7.3 vs. 5.2 months) | No significant difference<br>(11.8 vs. 10.1 months) | 48% vs. 38% | No significant difference<br>(4.5 vs. 3.2 months) | Not reported | No significant differences<br>in any subgroup |
| **Investigator 2** | No significant difference<br>(5.44 vs. 5.78 months) | No significant difference<br>(6.90 vs. 8.25 months) | 60.00% vs. 63.01% | No significant difference<br>(3.19 vs. 3.79 months) | Not reported | No significant differences<br>in any subgroup |
| **Investigator 3** | No significant difference<br>(4.67 vs. 5.14 months) | No significant difference<br>(9.17 vs. 9.25 months) | 0% vs. 0% | No significant difference<br>(3.19 vs. 3.79 months) | Not reported | No significant differences<br>in any subgroup |
| **Investigator 4** | No significant difference<br>(4.90 vs. 5.32 months) | No significant difference<br>(9.17 vs. 9.25 months) | 83.87% vs. 90.41% | Not reported | Not reported | Not reported |

#### 3.2.5 Survival Curves

One of the investigators generated Kaplan-Meier curves for progression-free survival (PFS) and overall survival (OS) using code produced by a Large Language Model (LLM) through specific prompting. The original publication and the investigator’s analyses showed no significant visual difference in the PFS and OS curves between the LY2510924 plus carboplatin/etoposide arm and the carboplatin/etoposide arm alone. The curves in both analyses underscored the aggressive nature of extensive-stage SCLC, with a rapid decline in survival probability in both treatment groups. The flattening tails of the curves in the investigator’s analysis highlighted the heterogeneity in patient outcomes and the presence of a subset of patients who experienced longer PFS and OS, regardless of the treatment arm. While visually similar, it is important to note that the survival curves generated by the LLM may not be directly comparable to those in the original publication due to potential differences in the underlying data and statistical methods used. Figure S1 (in section 2.0 of supplementary data) displays the Kaplan-Meier curves for progression-free survival (PFS) and overall survival (OS) in both treatment arms.

### 3.3 Analysis of Drug Safety

To thoroughly evaluate the safety profile of LY2510924 in combination with carboplatin/etoposide for the treatment of extensive-stage small cell lung cancer (ES-SCLC), four independent investigators conducted detailed analyses of adverse events (AEs) reported during the clinical trial. Their primary goal was to compare the incidence and severity of AEs in patients receiving LY2510924 plus carboplatin/etoposide (Arm A) versus those receiving carboplatin/etoposide alone (Arm B).

The original publication of the clinical trial data indicated that the overall safety results were similar between the two treatment arms. However, it also noted that some adverse events were more frequent in the LY2510924 plus carboplatin/etoposide group, suggesting a potential for increased toxicity with the combination therapy.

Independent investigators scrutinized the AE data to gain a deeper understanding of the safety profile. These analyses yielded mixed results, with some investigators finding no significant increase in AEs with the addition of LY2510924. In contrast, others observed a higher incidence of serious adverse events (SAEs) and treatment discontinuations due to AEs in Arm A. This variability in findings may be attributed to differences in the LLMs’ ability to identify and categorize adverse events and variations in the investigators’ prompting strategies when querying the models about safety data.

Across the analyses, the most common AEs were generally consistent and included neutropenia, nausea, fatigue, anemia, and thrombocytopenia. These are commonly associated with chemotherapy regimens, and their occurrence in both arms highlights the inherent challenges in managing side effects in ES-SCLC treatment.

#### 3.3.1 Notable Findings

Investigator 2’s analysis revealed a notably higher proportion of patients with SAEs in Arm A (51.06%) compared to Arm B (28.57%). Furthermore, more patients in Arm A (17.02%) discontinued treatment due to AEs compared to Arm B (4.76%). This suggests that adding LY2510924 may indeed increase the risk of serious side effects and lead to a higher treatment discontinuation rate. However, it is crucial to consider that these findings are based on the analysis of a single investigator and may not be fully representative.

Interestingly, the most common SAEs differed between the two arms. In Arm B (carboplatin/etoposide alone), the most frequent SAEs were pleural effusion and ataxia. In contrast, Arm A (LY2510924 combination) experienced more pneumonia, febrile neutropenia, and sepsis. This difference in SAE profiles could indicate that LY2510924 may contribute to specific types of serious adverse events.

*Table 3 summarizes the findings of the original publication and the four investigators’ analysis of adverse events*.

**Table 3.** Frequency and Severity of Adverse Events in the LY2510924 Clinical Trial.

| Study | Total AEs | Total SAEs | Discontinuations Due to AEs | Deaths | Most Common AEs | Most Common SAEs |
| --- | --- | --- | --- | --- | --- | --- |
| **Original Publication** | Not reported | Not reported | 17.0% (Arm A) vs. 4.7% (Arm B) | Not reported | Anemia, neutropenia, leukopenia, vomiting, pneumonia | Not reported |
| **Investigator 1** | 80% (Arm A) vs. 75% (Arm B) | 15% (Arm A) vs. 12% (Arm B) | 10% (Arm A) vs. 8% (Arm B) | 2% (Arm A) vs. 1% (Arm B) | Neutropenia, nausea, fatigue | Febrile neutropenia, pneumonia |
| **Investigator 2** | 100% (both arms) | 51.06% (Arm A) vs. 28.57% (Arm B) | 17.02% (Arm A) vs. 4.76% (Arm B) | Not reported | Anemia, fatigue, thrombocytopenia, neutropenia, nausea | Pleural effusion, ataxia (Arm B); Pneumonia, febrile neutropenia, sepsis (Arm A) |
| **Investigator 3** | 80% (Arm A) vs. 75% (Arm B) | 15% (Arm A) vs. 12% (Arm B) | 10% (Arm A) vs. 8% (Arm B) | 2% (Arm A) vs. 1% (Arm B) | Not reported | Not reported |
| **Investigator 4** | Not reported | Not reported | Not reported | Not reported | Anemia, fatigue, thrombocytopenia, nausea, neutropenia | Not reported |

### 3.4 Dropout Analysis

An analysis of patient dropouts was performed to assess the impact of treatment discontinuation on the study findings. *Table 4 summarizes the findings of Dropout Analysis by the four investigators*.

**Table 4.** Dropout Analysis.

| **Investigator** | **Total Enrolled** | **Completed Study** | **Dropouts** | **Reasons for Dropout** |
| --- | --- | --- | --- | --- |
| **Investigator**<br>**1** | 180 | 145 | 35 | Adverse events, disease progression, patient decision, non-compliance |
| **Investigator**<br>**2** | 89 | 74 | 15 | Adverse events, clinical progression, patient decision, non-compliance |
| **Investigator**<br>**3** | 92 | 55 | 37 | RECIST progression, adverse events, patient withdrawal of consent |
| **Investigator**<br>**4** | Not reported | Not reported | Not reported | Not reported |

The original publication did not report specific dropout numbers or reasons. However, it mentioned that 17.0% of patients in the LY2510924 plus carboplatin/etoposide arm discontinued treatment compared to 4.7% in the carboplatin/etoposide arm.

- **Investigator 1:** The majority of patients completed the study in both treatment groups, with slightly higher completion in the carboplatin/etoposide arm. Adverse events were the most common reason for dropout, with a slightly higher incidence in the LY2510924 combination arm.
- **Investigator 2:** Adverse events were the most common reason for dropout, suggesting that tolerability could be a limiting factor, particularly in the LY2510924 combination arm.
- **Investigator 3:** Approximately 59.8% of participants completed the study. The most common reasons for dropout were RECIST progression and adverse events.
- **Investigator 4:** No dropout analysis was reported by this investigator.

Despite some variations in reporting, the investigators’ analyses generally indicate that adverse events were a significant contributor to treatment discontinuation, particularly in the LY2510924 combination arm. This highlights the potential for increased toxicity with the addition of LY2510924. Disease progression was also a common reason for dropout, reflecting the aggressive nature of ES-SCLC.

#### Population Differences Among Investigators

An important observation is that the total number of enrolled participants reported by each investigator varies notably (Investigator 1: 180 participants; Investigator 2: 89 participants; Investigator 3: 92 participants; Investigator 4: not reported). This variation indicates that the investigators may have characterized different populations within the same dataset.

Possible reasons for these differences include:

- **Data Interpretation Variability:** The LLMs may have interpreted the inclusion criteria or data structures differently, leading to the selection of different cohorts.
- **Handling of Missing Data:** Each investigator might have employed different strategies for dealing with missing or imputed data, affecting the total participant count.

These differences in population characterization could contribute to the discrepancies noted in the Dropout Analysis and potentially impact other analyses outcomes.

In summary, the lack of detailed dropout information in the original publication and the variability in reporting across investigators limit the conclusions that can be drawn from this comparative analysis.

### 3.5 Biomarker Analysis

Both the original publication and one of the investigator’s analysis indicate that the addition of LY2510924 to carboplatin and etoposide does not result in significant changes in biomarker profiles. The original study reported that LY2510924 did not significantly alter CXCR4 expression in tumor cells but did show an increase in absolute CD34+ stem cell counts post-treatment in Arm A, confirming a pharmacodynamic response to CXCR4-SDF-1 axis inhibition. The investigator’s analysis was consistent with these findings. This consistency between the original publication and the LLM-driven analysis suggests that LLMs can effectively extract and interpret biomarker data from clinical trials.

Both analyses acknowledged the limitations posed by the small number and scope of biomarkers assessed, emphasizing the need for further research. Expanding the biomarker panel is crucial to fully understanding the biological effects of LY2510924 and identifying potential predictive biomarkers for treatment response.

## 4.0 DISCUSSION

This study explored the feasibility of using large language models (LLMs), specifically OpenAI’s ChatGPT-4 and Google’s Gemini Advanced, to analyze clinical trial data without specialized machine learning training. By employing a chain-of-thought (CoT) prompting template, we aimed to standardize the analytical approach and assess whether LLMs could produce results comparable to those achieved by experienced scientists.

### 4.1 Efficacy of LLMs in Clinical Data Analysis

The use of LLMs in analyzing the clinical trial data of LY2510924 combined with carboplatin and etoposide for extensive-stage small cell lung cancer (ES-SCLC) demonstrated both potential and limitations. Our findings reveal a mixed picture, highlighting both the promise and challenges of applying LLMs in this context. The LLMs were able to process complex datasets and generate analyses on baseline characteristics, efficacy endpoints, safety profiles, and biomarker data. This suggests that LLMs can serve as valuable tools in clinical research, potentially reducing the barriers associated with data interpretation for clinicians and researchers without advanced data science expertise.

### 4.2 Variability Between LLMs and Investigators

Despite using the same CoT prompting template and datasets, discrepancies were observed in the analyses produced by different investigators and LLMs. For instance, one investigator reported a statistically significant improvement in progression-free survival (PFS) with the addition of LY2510924, contradicting the original publication and the findings of other investigators.

Similarly, variations were noted in the analysis of objective response rate (ORR) and safety profiles.

These inconsistencies can be attributed to several factors:

- **Differences in Population Characterization:** Variations in how each investigator, guided by their respective LLMs, defined and selected the study population could lead to discrepancies in the analyses. This includes differences in handling missing data, applying inclusion/exclusion criteria, and interpreting data definitions.
- **Data Preprocessing Approaches:** Inconsistent preprocessing methods, such as data cleaning, normalization, and handling of outliers, can result in divergent datasets being analyzed, further contributing to variability in results.
- **Model Drift:** Updates to LLMs over time can lead to changes in their behavior, known as model drift. If investigators used different versions of the LLMs or accessed them at different times, the outputs could vary due to updates in the models’ parameters or training data.
- **Differences in LLM Capabilities:** While both ChatGPT-4 and Gemini Advanced are advanced LLMs, they may have differing architectures, training data, and inherent biases that affect their analytical outputs.
- **Prompting Strategies:** Minor variations in how investigators phrased their prompts could lead to different interpretations by the LLMs. The models are sensitive to input phrasing, and subtle differences can significantly impact the output.
- **Data Interpretation Limitations:** LLMs may struggle with interpreting complex statistical data, especially if the data requires nuanced understanding beyond text-based information.
- **Emergent Properties and Generative AI:** As generative AI models, LLMs exhibit emergent properties, complex behaviors that arise unpredictably and cannot be fully explained by their underlying architecture or training data. This inherent unpredictability can introduce variability in their outputs, potentially leading to inconsistencies in analysis outcomes, even when the models are provided with identical data and prompts.

### 4.3 Impact of Chain-of-Thought Prompting

Our CoT prompting template was designed to guide the LLMs through a structured analytical process, mirroring the reasoning of experienced data scientists. By providing explicit step-by-step instructions and emphasizing critical aspects of the analysis, we aimed to standardize outputs across different models and investigators. While this approach demonstrated value in organizing the analysis, the observed variability in our results suggests that current LLM models still require careful oversight to ensure accuracy and consistency.

Recent advances in LLM architectures, such as GPT-4 o1, have incorporated intrinsic CoT reasoning, allowing models to execute multi-step logical processes without requiring explicitly engineered prompts. However, GPT-4 o1 was not available at the time of our analysis, and its capacity for clinical trial data interpretation has yet to be systematically evaluated. Other advanced models, such as Claude and Gemini, have since integrated native CoT capabilities, which may reduce prompt dependency and improve output stability. Future research should assess whether these models, with inherently structured reasoning processes, can yield more reproducible and robust results when applied to clinical trial data.

The CoT process is dependent on the model’s ability to follow logical reasoning, which may not always align with human analytical methods. Incorporating a specialized CoT framework tailored to clinical trial analysis could bridge this gap by providing more explicit guidance on interpreting clinical data nuances. Additionally, given the emergent properties of LLMs, ensuring consistent and accurate interpretation of complex clinical trial data remains a challenge. To enhance the applicability of LLMs for clinical trial analysis, future research should focus on developing alignment strategies that optimize the model’s reasoning capabilities to address the complexities of clinical data. This may include adapting CoT frameworks to better process or interpret factors such as treatment effects, patient demographics, statistical methodologies (e.g., survival analysis), and interdependencies in key clinical variables. Achieving alignment with these clinical and statistical considerations will be critical to improving the consistency, accuracy, and reliability of LLM-driven analyses in clinical trial settings. Further investigations could also explore the integration of LLMs with complementary analytical tools to strengthen the robustness and reproducibility of the analyses.

### 4.4 Differences Between Original and Current Analysis

A significant factor influencing the discrepancies between our findings and the original publication is the difference in the underlying datasets. The data retrieved from the Project Data Sphere (PDS) platform were de-identified and modified, with missing data points and imputed values, compared to the original study data. Many data points were imputed or removed to prevent re-analysis of the clinical trial data, which could lead to limitations in the depth and accuracy of the analysis. These limitations may have contributed to the inconsistencies observed in the analyses and highlight the importance of data quality and completeness for reliable LLM-driven analysis.

### 4.6 Potential and Future Applications

Despite the limitations, our study demonstrates the potential utility of current LLMs in clinical trial data analysis. With further refinement, including integration with statistical software and enhancements in data interpretation capabilities, LLMs could become valuable tools for clinicians and researchers. They could facilitate quicker preliminary analyses, generate hypotheses, and assist in drafting reports, thereby accelerating the research process.

A particularly promising direction involves the implementation of specialized CoT frameworks optimized for clinical trial analysis. These frameworks could leverage the semantic processing capabilities inherent in contemporary LLM architectures while maintaining analytical rigor. The incorporation of domain-specific knowledge and structured analytical protocols into CoT prompting mechanisms may enhance the models’ capacity for consistent and accurate data interpretation.

### 4.7 Recommendations for Improved Integration

To enhance the effectiveness of LLMs in clinical data analysis:

- **Hybrid Approaches:** Combining LLMs with validated statistical software platforms could mitigate computational limitations, allowing for accurate statistical analyses guided by LLM-generated insights.
- **Enhanced Prompting Techniques:** Development and standardization of precise prompting methodologies may enhance output consistency and reproducibility. Systematic training in prompt engineering techniques constitutes a critical component of successful implementation, requiring standardized protocols and ongoing evaluation.
- **Data Quality Assurance:** The accuracy and completeness of input data directly determines the reliability of LLM-generated analyses, necessitating comprehensive quality control measures throughout the analytical pipeline.
- **Consideration of LLM Characteristics:** Future research should include comparative analyses of various LLM architectures’ performance across different clinical trial data types and analytical tasks. This evaluation must account for models’ emergent properties, generative characteristics, and inherent limitations in processing numerical and statistical information.
- **Tailored CoT Frameworks:** Creation of specialized CoT frameworks incorporating domain-specific logical sequences and analytical considerations can enhance analytical precision. These frameworks should be designed to guide LLMs through complex clinical data analysis while maintaining methodological rigor and reproducibility.

## 5.0 CONCLUSIONS AND FUTURE DIRECTIONS

This study examined the use of the native capabilities of LLMs, specifically ChatGPT-4 and Gemini Advanced, in analyzing clinical trial data. While the LLMs demonstrated the ability to process and interpret complex clinical data, inconsistencies in their outputs highlight current limitations in their application for rigorous and reproducible data analysis.

Our implementation of CoT prompting established a structured analytical framework designed to enhance methodological consistency. However, our analysis identified several critical limiting factors: variations in LLM capabilities, inconsistencies in prompting effectiveness, and dataset completeness constraints. These limitations impacted both the reproducibility of analyses and our ability to validate results against the original study findings.

Despite the identified limitations, LLMs hold significant potential for democratizing and accelerating clinical trial data analysis, potentially broadening access for clinicians and researchers without advanced data science and statistical expertise. While LLMs cannot yet replace human experts, they can provide valuable insights to assist clinicians and researchers in interpreting complex clinical trial data.

Future development priorities for LLMs in clinical trial analysis must address several interconnected challenges to enhance their practical implementation and reliability. The optimization of statistical computation capabilities is essential for improving accuracy in complex analyses, with particular emphasis on validating numerical calculations and statistical inference methodologies. This technical foundation requires robust data quality management protocols that ensure consistent analytical inputs across diverse clinical trial datasets, incorporating standardized preprocessing, validation checks, and error detection mechanisms. The refinement of specialized CoT frameworks, optimized for clinical trial analysis, could substantially improve both consistency and reproducibility across different trial contexts. Such frameworks could enable automated reporting of clinical trial results, supporting interim evaluations [18, 19] without unblinding patient-level data, while facilitating ongoing recruitment analysis and adaptive randomization [20, 21, 22, 23, 24, 25, 26].

To realize the full potential of LLMs in clinical research, future investigations should systematically examine fundamental model characteristics, analytical capabilities, and computational boundaries—with particular attention to their increasingly emergent properties and generative behaviors. This requires rigorous validation against established methodological standards, while carefully examining the interplay between model architecture, computational constraints, and analytical reliability. By investigating diverse therapeutic applications and trial designs, we can develop a comprehensive understanding of the factors that influence model performance to unlock their transformative potential. Through these coordinated advances, LLMs have the potential to fundamentally transform clinical trial execution and analysis— accelerating drug development, improving trial design, and ultimately expediting the delivery of novel therapeutics to patients while maintaining the highest standards of scientific rigor.

## 6.0 LIMITATIONS

Several limitations warrant examination in our study, particularly regarding the intersection of model capabilities, methodological constraints, and data integrity. These limitations provide important context for understanding both the current constraints and future potential of LLMs in clinical trial analysis.

### Model Evolution and Performance Considerations

The inconsistencies observed across investigator analyses, particularly in endpoint interpretation and statistical calculations, suggest potential impacts from model evolution over time. Large language models (LLMs) are continuously updated, with modifications to their architectures, training data, and optimization parameters. These updates, while improving overall performance, can introduce variability in analytical outputs—a phenomenon known as **model drift** (Bommasani et al., 2021) [12]. Model drift can manifest as changes in statistical reasoning, different interpretations of similar prompts, or shifts in numerical processing capabilities. If different investigators accessed the LLMs at different times, disparities in results may stem from such temporal variations in model behavior. Ensuring version control and maintaining detailed documentation on model updates will be critical for the reproducibility of LLM-driven analyses.

### Computational and Analytical Constraints

Our findings indicate that while LLMs can perform basic statistical reasoning, they exhibit limitations in handling complex numerical analyses with precision. This was particularly evident in survival analyses and objective response rate (ORR) calculations, where discrepancies across investigators suggest that LLMs may struggle with maintaining consistency in statistical outputs. Unlike traditional biostatistical tools, LLMs are not designed for high-precision statistical modeling, and their results require verification through established methodologies (Titus, 2023) [10]. This limitation underscores the need for hybrid approaches that integrate LLMs with specialized statistical software to ensure analytical accuracy.

### Data Quality and Impact on Results

The study utilized modified clinical trial data from the Project Data Sphere platform, which had undergone de-identification and imputation (Green et al., 2015) [16]. These modifications, while necessary for data privacy, may have introduced inconsistencies that affected the validity of the analyses. Variations in cohort sizes reported by investigators (ranging from 89 to 180 participants) suggest differences in how the LLMs processed missing data, applied inclusion/exclusion criteria, or interpreted dataset structures. These discrepancies highlight the need for standardized data preprocessing and interpretation frameworks to minimize variability in LLM-driven analyses.

### Framework and Validation Constraints

Although the **Chain-of-Thought (CoT) prompting framework** was designed to provide structural guidance for LLM analyses, inconsistencies in outputs suggest that it may not yet be optimized for clinical trial data interpretation (Wei et al., 2022) [14]. The variability observed in endpoint analyses, safety profiles, and biomarker assessments indicates that LLMs require further refinement to reliably handle the complexity of clinical trial datasets. The absence of external statistical verification in this study also limits the ability to assess the full accuracy of

LLM-generated results. Future studies should incorporate additional validation steps, including comparisons against independent biostatistical analyses, to strengthen the credibility of LLM-driven insights.

### Methodological and Investigator Variability

The study relied on four independent investigators using LLMs to conduct their analyses, and while efforts were made to standardize prompts and methodologies, human variability in prompt phrasing and data interpretation likely contributed to inconsistencies in the results. LLMs are highly sensitive to subtle variations in input, which can lead to divergent outputs even when analyzing the same dataset (Lee et al., 2023) [6]. The introduction of more structured and standardized analytical frameworks may help mitigate this issue in future applications.

### Future Research Considerations

To improve the reliability of LLMs in clinical trial analysis, several key areas require further investigation:

- **Standardized Data Handling Protocols**: Establishing clear guidelines for dataset preprocessing, missing data imputation, and cohort selection will help ensure consistency in LLM-driven analyses.
- **Version Control for LLMs**: Documenting model versions used in research and assessing the impact of model drift on analytical outputs will be essential for reproducibility.
- **Integration with Statistical Software**: Combining LLMs with validated biostatistical tools can enhance analytical accuracy and provide a hybrid approach that balances automation with precision.
- **Enhanced CoT Prompting Strategies**: Developing domain-specific CoT frameworks tailored to clinical trial analysis could improve consistency and reduce variability in LLM-generated results.

Understanding these limitations provides crucial context for future research while highlighting the importance of careful methodology in evaluating LLM capabilities for clinical trial analysis. By addressing these constraints, future investigations can establish more robust frameworks for integrating LLMs into clinical research while maintaining scientific rigor.

## Supporting information

Supplementary 2 - Investigator1 Report

Supplementary 3 - Investigator2 Report

Supplementary 4 - Investigator3 Report

Supplementary 5 - Investigator4 Report

## Data Availability

The data generated in this study are provided as supplementary materials. The de-identified patient-level data from the clinical trial used in this analysis is available at Project Data Sphere Platform (https://data.projectdatasphere.org/). Access to this data can be requested through the platform.

https://data.projectdatasphere.org/

## Acknowledgments and Funding

Dr. Komandur, Dr. McDunn and Dr. Khozin acknowledges funding support from CEO Roundtable on Cancer and Project Data Sphere.

Dr. Dicker is employed by Thomas Jefferson University/Jefferson Health. His work receives additional support from the National Cancer Institute, NRG Oncology, and the Prostate Cancer Foundation Challenge Grant.

