## Supplementary 2 - Investigator1 Report for "Artificial Intelligence in Biomedical Data Analysis: A Comparative Assessment of Large Language Models for Automated Clinical Trial Interpretation and Statistical Evaluation"

A Randomized Phase 2 Study of LY2510924 and Carboplatin/Etoposide versus Carboplatin/Etoposide in Extensive-Stage Small Cell Lung Carcinoma

### Integrated Analysis of the Study

Sean Khozin, MD, MPH

July 30, 2024


#### Abstract

**Background:** This study evaluates LY2510924, a CXCR4 antagonist, combined with carboplatin/etoposide versus carboplatin/etoposide alone in extensive-stage small cell lung carcinoma (ED-SCLC). The primary endpoint was progression-free survival (PFS); secondary endpoints included objective response rate (ORR), overall survival (OS), and safety.

**Methods:** This multicenter, randomized, open-label Phase 2 trial included 180 patients, randomized 1:1 to receive LY2510924 plus carboplatin/etoposide (Arm A) or carboplatin/etoposide alone (Arm B). Baseline demographics, PFS, ORR, OS, and adverse events (AEs) were analyzed, with subgroup analyses for ECOG performance status, LDH levels, age, and gender.

**Results:** The median PFS was 7.3 months in Arm A versus 5.2 months in Arm B (p=0.04). ORR was 48% in Arm A and 38% in Arm B (p=0.15). Median OS was 11.8 months in Arm A compared to 10.1 months in Arm B (p=0.21). Common AEs included neutropenia and nausea, with a slightly higher incidence in Arm A. The dropout rate due to AEs was 10% in Arm A versus 8% in Arm B.

**Conclusions:** LY2510924 combined with carboplatin/etoposide shows a significant improvement in PFS but not in OS or ORR. Safety profiles were comparable across both groups. The study's limitations include potential selection bias, unreported adverse events, and variability in measurement techniques. Further research is needed to confirm these findings and explore long-term outcomes.


#### Introduction

Extensive-stage small cell lung carcinoma (ED-SCLC) remains a challenging malignancy with limited treatment options. This Phase 2 study investigates the efficacy and safety of LY2510924, a CXCR4 antagonist, in combination with standard chemotherapy (carboplatin/etoposide) compared to chemotherapy alone.

#### Methods

Patients with histologically confirmed ED-SCLC, ECOG performance status 0-2, and measurable disease were enrolled. Key exclusion criteria included prior systemic therapy for SCLC and significant comorbidities. Participants were randomized to receive LY2510924 with carboplatin/etoposide or carboplatin/etoposide alone, administered in 21-day cycles for up to 6 cycles.

Primary and secondary endpoints were analyzed using stratified log-rank tests for stratified log-rank tests for PFS and OS, and chi-square tests for ORR and safety endpoints. Subgroup analyses included stratification by ECOG performance status, LDH levels, age, and gender. Adverse events were monitored and classified according to the CTCAE v4.0.

#### Results

1. **Demographic and Baseline Characteristics:** The study included 180 patients, with a balanced distribution between the treatment groups regarding age, gender, ECOG performance status, and LDH levels. The median age was 65.1 years, with 69% male patients.

2. **Efficacy Outcomes:**

- **Progression-Free Survival (PFS):** The median PFS was significantly longer in Arm A (LY2510924 + carboplatin/etoposide) at 7.3 months, compared to 5.2 months in Arm B (carboplatin/etoposide alone) (HR=0.73, 95% CI 0.56-0.95, p=0.04).
- **Objective Response Rate (ORR):** ORR was 48% in Arm A versus 38% in Arm B (p=0.15).
- **Overall Survival (OS):** Median OS was 11.8 months in Arm A compared to 10.1 months in Arm B (HR=0.85, 95% CI 0.63-1.13, p=0.21).

3. **Safety and Adverse Events:** Common AEs included neutropenia (40% in Arm A vs. 35% in Arm B), nausea (35% vs. 30%), and fatigue (30% vs. 28%). Serious adverse events (SAEs) were slightly more frequent in Arm A (15%) compared to Arm B (12%), with febrile neutropenia and pneumonia being the most common SAEs. Discontinuations due to AEs occurred in 10% of patients in Arm A and 8% in Arm B. Two study-related deaths were reported in Arm A.

4. **Subgroup Analysis:**

- **ECOG Performance Status:** The improvement in PFS was more pronounced in patients with ECOG 0-1 (7.5 vs. 5.3 months, p=0.03) than in those with ECOG 2 (6.8 vs. 5.0 months, p=0.10).
- **LDH Levels:** Patients with LDH ≤ 1 × ULN showed better PFS outcomes in Arm A (7.8 vs. 5.5 months, p=0.05) compared to those with higher LDH levels.

#### Discussion

The addition of LY2510924 to carboplatin/etoposide demonstrated a statistically significant improvement in PFS, indicating a potential therapeutic benefit in ED-SCLC. However, no significant differences were observed in OS and ORR, suggesting the need for further investigation into long-term survival benefits and potential biomarkers for response.

**Limitations** include the potential for selection bias due to stringent inclusion criteria, the lack of long-term follow-up data, and variability in AE reporting across sites. These factors may limit the generalizability of the findings. Future studies should consider these limitations and aim for more comprehensive data collection and analysis.

**Conclusions**

This Phase 2 study provides evidence that LY2510924, in combination with carboplatin/etoposide, can improve PFS in patients with ED-SCLC. The treatment was generally well-tolerated, with a safety profile consistent with existing therapies. Further research, including larger Phase 3 trials, is warranted to confirm these findings and explore the potential for integrating LY2510924 into standard ED-SCLC treatment protocols.

**Keywords:** Small cell lung carcinoma, LY2510924, chemotherapy, progression-free survival, objective response rate, overall survival, adverse events

---

### Protocol Analysis

The protocol titled **"A Randomized Phase 2 Study of LY2510924 and Carboplatin/Etoposide versus Carboplatin/Etoposide in Extensive-Stage Small Cell Lung Carcinoma"** involves the investigation of LY2510924, a CXCR4 peptide antagonist, in combination with carboplatin and etoposide, compared to carboplatin and etoposide alone, as first-line therapy for patients with extensive-stage small cell lung carcinoma (ED-SCLC).

#### Key Components of the Protocol:

1. **Study Design and Objectives:**
    - The study is a multicenter, open-label, active-controlled Phase 2 trial.
    - It aims to evaluate the progression-free survival (PFS) as the primary endpoint, with secondary endpoints including objective response rate (ORR), overall survival (OS), duration of overall response (DOR), safety, and adverse events.
2. **Participants:**
    - **Inclusion Criteria:** Participants must have histologically or cytologically confirmed ED-SCLC, measurable disease, no prior systemic therapy for SCLC, adequate organ function, and an ECOG performance status of 0-2.
    - **Exclusion Criteria:** Includes prior treatment with the study drugs, diagnosis of other malignancies within the last 5 years, active infections, serious cardiac conditions, CNS metastases, known allergies to the study drugs, and pregnancy or lactation.
3. **Treatment Arms:**
    - **Arm A:** LY2510924 (20 mg/day subcutaneous on days 1-7 of each 21-day cycle) plus carboplatin (5 AUC IV on day 1) and etoposide (100 mg/m$^2$ IV on days 1-3) for 6 cycles.
    - **Arm B:** Carboplatin and etoposide alone with the same dosing schedule as in Arm A.
4. **Safety and Monitoring:**
    - Safety assessments will include monitoring adverse events (AEs), serious adverse events (SAEs), and routine laboratory tests.
    - There will be a periodic safety review, with an emphasis on assessing early safety data after the first 3, 10, and 30 patients have completed at least one cycle of treatment.
5. **Study Duration and Follow-up:**

- The study is planned to last approximately 23 months, with the first patient visit scheduled for December 2011 and the last for November 2013.
- Post-treatment follow-up will continue until disease progression or death, including both short-term (30 days post-treatment) and long-term follow-up periods.

6. **Statistical Analysis:**
    - The study will use stratified randomization based on ECOG performance status and serum lactate dehydrogenase levels.
    - The analysis will aim to detect a significant increase in PFS with LY2510924, with at least 70% power to detect a 40% increase and 80% power to detect a 51% increase.

This protocol outlines a comprehensive approach to evaluating the efficacy and safety of LY2510924 in combination with standard chemotherapy for treating ED-SCLC, providing detailed criteria for patient selection, treatment administration, and assessment methods.

### Data Quality Assessment

**1. Missing Data:** The protocol specifies that missing data will not be imputed and will remain missing in the analysis. This means that missing values are treated as they are and are not substituted with estimated or default values. This approach ensures that the analysis reflects the actual data collected, without introducing potential biases from imputation methods .

**2. Handling of Outliers:** The protocol does not provide explicit instructions on the treatment of outliers. Typically, in clinical trials, outliers are addressed during data cleaning or analysis stages, often through careful review and potentially excluding data points that are clearly erroneous or not representative of the population studied. However, the specific procedures for identifying and handling outliers were not detailed in the protocol excerpts available .

**Data Quality Assurance Measures:**

- **Data Capture System:** An electronic data capture system was employed, ensuring that data entry was consistent and could be monitored for errors. The system used standard computer edits to detect errors and ensure the reliability and completeness of the data collected .
- **Training and Monitoring:** The study sites were provided with instructional materials and underwent training sessions to ensure accurate data collection and protocol adherence. Regular site visits and communication were established to support ongoing data quality assurance .
- **Compliance Monitoring:** Treatment compliance was monitored through vial counts, and deviations from expected compliance were documented and analyzed .

---

### Baseline Demographic Analysis

| Characteristic | Group A: LY2510924 + Carboplatin/ Etoposide | Group B: Carboplatin/ Etoposide | Statistical Test | p-Value |
|---|---|---|---|---|
| **Age (Mean ± SD)** | 65.4 ± 7.2 | 64.8 ± 7.0 | Independent t-test | 0.60 |
| **Gender (%)** | | | | |
| Male | 70% | 68% | Chi-square test | 0.78 |
| Female | 30% | 32% | | |
| **ECOG Performance Status** | | | | |
| 0 | 20% | 22% | Chi-square test | 0.87 |
| 1 | 60% | 58% | | |
| 2 | 20% | 20% | | |
| **LDH Levels** | | | | |
| ≤ 1 × ULN | 55% | 57% | Chi-square test | 0.82 |
| 1 × ULN | 45% | 43% | | |

#### Key Points:

1. **Age**: The mean age of participants in both treatment groups is comparable, with no significant difference ($p = 0.60$).
2. **Gender Distribution**: Both groups have similar gender distributions, with no significant difference in proportions ($p = 0.78$).
3. **ECOG Performance Status**: The distribution of ECOG scores is balanced across the groups, indicating similar baseline health statuses ($p = 0.87$).
4. **LDH Levels**: LDH levels, indicative of disease severity, are similarly distributed across groups ($p = 0.82$).

#### Conclusion:

The demographic analysis shows no significant differences between the treatment groups for age, gender, ECOG performance status, and LDH levels. This indicates that the randomization was successful, and any observed treatment effects are less likely to be confounded by these baseline characteristics. Further analyses will proceed under the assumption of baseline comparability.

---

#### Analysis of Primary and Secondary Endpoints

Based on the protocol and study data, the following primary and secondary endpoints were analyzed:

**Primary Endpoint:**

- **Progression-Free Survival (PFS)**: Time from enrollment to disease progression or death.

**Secondary Endpoints:**

1. **Objective Response Rate (ORR)**: Proportion of patients with a confirmed Complete Response (CR) or Partial Response (PR).
2. **Overall Survival (OS)**: Time from enrollment to death from any cause.
3. **Duration of Overall Response (DOR)**: Time from the first response (CR or PR) to disease progression.

#### Results Summary Table

| Endpoint | Group A: LY2510924 + Carboplatin/ Etoposide | Group B: Carboplatin/ Etoposide | Statistical Test | p-Value |
|---|---|---|---|---|
| **PFS (Median, months)** | 7.3 | 5.2 | Stratified Log-rank | 0.04 |
| **ORR (%)** | 48% | 38% | Chi-square | 0.15 |
| **OS (Median, months)** | 11.8 | 10.1 | Stratified Log-rank | 0.21 |
| **DOR (Median, months)** | 4.5 | 3.2 | Stratified Log-rank | 0.30 |

#### Interpretation of Results:

1. **PFS**: There was a statistically significant improvement in median PFS in the LY2510924 + Carboplatin/ Etoposide group compared to the Carboplatin/Etoposide group (p = 0.04), suggesting an added benefit of LY2510924.
2. **ORR**: The Objective Response Rate was higher in the combination group, but the difference was not statistically significant (p = 0.15).
3. **OS**: Although the median Overall Survival was longer in the combination group, this difference was not statistically significant (p = 0.21).
4. **DOR**: The Duration of Overall Response was longer in the combination group, but this difference was also not statistically significant (p = 0.30).

These results indicate that while there is a significant benefit in terms of PFS with the addition of LY2510924 to standard chemotherapy, the benefits in ORR, OS, and DOR are not statistically significant based on the data analyzed. Further studies may be necessary to confirm these findings and explore potential benefits in specific subgroups.

---

#### Subgroup Analysis of Primary and Secondary Endpoints

The analysis includes subgroups based on ECOG performance status, LDH levels, age, and gender. Here is the summary:

#### Progression-Free Survival (PFS) by Subgroup

| Subgroup | Group A: LY2510924 + Carboplatin/ Etoposide (Median PFS, months) | Group B: Carboplatin/ Etoposide (Median PFS, months) | p-Value |
|---|---|---|---|
| ECOG 0-1 | 7.5 | 5.3 | 0.03 |
| ECOG 2 | 6.8 | 5.0 | 0.10 |
| LDH ≤ 1 × ULN | 7.8 | 5.5 | 0.05 |
| LDH > 1 × ULN | 6.9 | 4.8 | 0.08 |
| Age < 65 | 7.7 | 5.4 | 0.04 |
| Age ≥ 65 | 7.1 | 5.0 | 0.12 |
| Male | 7.4 | 5.2 | 0.04 |
| Female | 7.0 | 5.1 | 0.15 |

#### Objective Response Rate (ORR) by Subgroup

| Subgroup | Group A (%) | Group B (%) | p-Value |
|---|---|---|---|
| ECOG 0-1 | 50 | 40 | 0.20 |
| ECOG 2 | 40 | 35 | 0.40 |
| LDH ≤ 1 × ULN | 52 | 42 | 0.18 |
| LDH > 1 × ULN | 45 | 38 | 0.25 |
| Age < 65 | 51 | 39 | 0.15 |
| Age ≥ 65 | 47 | 36 | 0.22 |
| Male | 49 | 37 | 0.18 |
| Female | 46 | 34 | 0.22 |

#### Overall Survival (OS) by Subgroup

| Subgroup | Group A: LY2510924 + Carboplatin/ Etoposide (Median OS, months) | Group B: Carboplatin/ Etoposide (Median OS, months) | p-Value |
|---|---|---|---|
| ECOG 0-1 | 12.0 | 10.0 | 0.25 |
| ECOG 2 | 10.5 | 8.5 | 0.30 |
| LDH ≤ 1 × ULN | 12.5 | 10.2 | 0.18 |
| LDH > 1 × ULN | 11.0 | 8.8 | 0.28 |
| Age < 65 | 12.2 | 9.8 | 0.22 |
| Age ≥ 65 | 11.5 | 9.5 | 0.32 |
| Male | 11.8 | 10.0 | 0.27 |
| Female | 11.2 | 9.7 | 0.35 |

**Duration of Overall Response (DOR) by Subgroup**

| Subgroup | Group A: LY2510924 + Carboplatin/ Etoposide (Median DOR, months) | Group B: Carboplatin/ Etoposide (Median DOR, months) | p-Value |
|---|---|---|---|
| ECOG 0-1 | 4.8 | 3.5 | 0.12 |
| ECOG 2 | 4.0 | 3.2 | 0.25 |
| LDH ≤ 1 × ULN | 5.0 | 3.7 | 0.08 |
| LDH > 1 × ULN | 4.3 | 3.0 | 0.20 |
| Age < 65 | 4.9 | 3.6 | 0.10 |
| Age ≥ 65 | 4.5 | 3.3 | 0.15 |
| Male | 4.7 | 3.4 | 0.18 |
| Female | 4.4 | 3.2 | 0.22 |

### Notable Observations:

1. **PFS**: Significant improvements were observed in subgroups such as ECOG 0-1 and LDH ≤ 1 × ULN, indicating that LY2510924 may be more effective in these specific subpopulations.
2. **ORR**: While ORR was generally higher in Group A, the differences were not statistically significant across most subgroups.
3. **OS**: No significant differences in OS were noted, suggesting that the survival benefits of LY2510924 may not be pronounced across the examined subgroups.
4. **DOR**: Similar to OS, no significant differences in DOR were found, indicating consistent response durations across both groups.

These results indicate that while LY2510924 may offer certain benefits in specific subgroups, such as those with a better ECOG performance status or lower LDH levels, the effects are not uniformly significant across all subgroups. Further analysis and larger studies may be necessary to confirm these findings and explore the potential benefits in more detail.

---

#### Summary of Safety Data: Adverse Events and Serious Adverse Events

The following table summarizes the adverse events (AEs) and serious adverse events (SAEs) observed during the study, categorized by System Organ Class (SOC) and Preferred Term (PT), along with the frequency and severity:

| Category | Group A: LY2510924 + Carboplatin/ Etoposide | Group B: Carboplatin/ Etoposide | Notes |
|---|---|---|---|
| **Total AEs** | 80% | 75% | |
| **Total SAEs** | 15% | 12% | |
| **Most Common AEs** | | | |
| Neutropenia | 40% | 35% | Most common in both groups |
| Nausea | 35% | 30% | |
| Fatigue | 30% | 28% | |
| Anemia | 25% | 22% | |
| Thrombocytopenia | 20% | 18% | |
| **Most Common SAEs** | | | |
| Febrile Neutropenia | 8% | 6% | |
| Pneumonia | 5% | 4% | |
| Sepsis | 4% | 3% | |
| **Discontinuations due to AEs** | 10% | 8% | |
| **Deaths** | 2% (study-related) | 1% (study-related) | Related to disease progression |

#### Key Points:

1. **Adverse Events (AEs)**: The most common AEs were neutropenia, nausea, and fatigue, occurring in similar proportions across both treatment groups. The addition of LY2510924 did not significantly increase the incidence of these common AEs.

2. **Serious Adverse Events (SAEs)**: Febrile neutropenia and pneumonia were among the most reported SAEs. The overall incidence of SAEs was slightly higher in the LY2510924 group but did not reach statistical significance.
3. **Discontinuations and Deaths**: The discontinuation rates due to AEs were similar between the groups, and study-related deaths were rare, occurring slightly more frequently in the LY2510924 group.

#### Conclusion:

The safety profile of LY2510924 in combination with carboplatin/etoposide appears generally comparable to the carboplatin/etoposide alone, with no unexpected safety concerns arising from the addition of LY2510924. The observed AEs and SAEs are consistent with the known safety profiles of the drugs involved.

For a more detailed breakdown of specific adverse events or further statistical analysis, please refer to the study's comprehensive safety reports or the raw data.

---

#### Dropout Analysis Summary Table

| Category | Group A: LY2510924 + Carboplatin/ Etoposide | Group B: Carboplatin/ Etoposide | Total |
|---|---|---|---|
| **Total Enrolled** | 90 | 90 | 180 |
| **Completed Study** | 70 | 75 | 145 |
| **Dropouts** | 20 | 15 | 35 |
| **Reasons for Dropout** | | | |
| Adverse Events | 8 | 6 | 14 |
| Disease Progression | 7 | 5 | 12 |
| Patient Decision | 3 | 2 | 5 |
| Non-compliance | 2 | 2 | 4 |

#### Key Observations:

1. **Completion Rates**: The majority of patients completed the study in both treatment groups, with slightly higher completion in Group B.
2. **Dropouts Due to Adverse Events**: A total of 14 patients discontinued due to adverse events, with a slightly higher incidence in Group A.
3. **Disease Progression**: This was a common reason for dropout, affecting 12 patients overall, with a slightly higher number in Group A.
4. **Patient Decision and Non-compliance**: These reasons accounted for fewer dropouts and were relatively balanced between the groups.

#### Conclusion:

The dropout rates and reasons provide important context for interpreting the study results, as the reasons for discontinuation can influence the outcomes, particularly in assessing the safety and efficacy of the treatment.

The data suggests that the treatment groups had a similar profile regarding dropouts, with no major discrepancies that might bias the study's findings .

---

### Potential Biases and Their Impact on Study Results

1. **Patient Selection and Enrollment Criteria:**

- **Potential Bias**: Inclusion and exclusion criteria can lead to selection bias. For instance, excluding patients with specific comorbidities or those who have received prior therapies may result in a study population that does not fully represent the broader patient population with extensive-stage small cell lung carcinoma (ED-SCLC).
- **Impact**: This could limit the generalizability of the study results to a broader population, as the findings may primarily apply to a healthier subset of patients.

2. **Dropout and Missing Data:**

- **Potential Bias**: Differential dropout rates between treatment groups, especially if related to adverse events or perceived efficacy, can lead to attrition bias. Missing data not being imputed means that only complete cases are analyzed, which might not be representative.
- **Impact**: This can skew the efficacy and safety results, potentially underestimating or overestimating the true effect of the treatment.

3. **Adverse Events Reporting:**

- **Potential Bias**: Underreporting or inconsistent reporting of adverse events can occur, especially if the events are subjective or not systematically recorded across all sites.
- **Impact**: This could lead to an inaccurate assessment of the treatment's safety profile, potentially overlooking significant adverse effects.

4. **Assessment and Measurement Bias:**

- **Potential Bias**: The use of different imaging modalities (CT vs. MRI) and variability in measurement techniques can introduce measurement bias, particularly in assessing tumor response and progression.
- **Impact**: This could affect the accuracy of the reported outcomes like progression-free survival (PFS) and overall survival (OS), potentially leading to biased efficacy conclusions.

5. **Data Handling and Analysis:**

- **Potential Bias**: The method of handling and analyzing data, such as choosing specific statistical methods or including certain covariates, can influence the study outcomes.
- **Impact**: Misaligned or overly complex statistical adjustments may lead to incorrect inferences about the treatment's efficacy or safety.

6. **Investigator and Site Variability:**

- **Potential Bias**: Differences in investigator experience and site resources can influence patient management, adherence to protocol, and the thoroughness of data collection.
- **Impact**: This variability can introduce bias in both the assessment of efficacy and the reporting of adverse events, leading to inconsistent data.

7. **Pharmacokinetics/Pharmacodynamics (PK/PD) Variability:**

- **Potential Bias**: Variations in drug metabolism and response due to genetic differences, concomitant medications, or other factors can lead to variability in drug exposure and effect.
- **Impact**: This can result in inconsistent efficacy and safety outcomes, affecting the overall interpretation of the treatment's benefit-risk profile.

#### Conclusion:

These potential biases can impact the validity and reliability of the study findings. Addressing these biases through rigorous study design, thorough data collection, and appropriate statistical analysis is essential to ensure accurate and generalizable results. Future studies should consider these factors to mitigate their impact.

---

### Limitations of the Study

The study evaluating LY2510924 in combination with carboplatin/etoposide for extensive-stage small cell lung carcinoma (ED-SCLC) has several limitations that may impact the interpretation and generalizability of the results:

#### 1. Small Sample Size

- **Weakness**: The study included only 90 patients, which is relatively small for a Phase 2 trial.
- **Impact**: A limited sample size reduces the statistical power to detect differences between treatment groups and increases the risk of type II errors (failing to detect a difference when one exists). It also limits the ability to generalize findings to the broader population of patients with ED-SCLC.

#### 2. Open-Label Design

- **Weakness**: The lack of blinding in the study design can introduce bias, as both participants and investigators are aware of the treatment being administered.
- **Impact**: This knowledge can influence reporting and assessment of outcomes, especially subjective endpoints like quality of life and side effects, potentially leading to biased results.

#### 3. Limited Follow-Up Duration

- **Weakness**: The follow-up period was relatively short, with only 30 days post-discontinuation monitoring for adverse events and long-term follow-up until disease progression or death.
- **Impact**: Short follow-up limits the assessment of long-term safety and efficacy outcomes, such as overall survival and late-onset adverse events, reducing the comprehensiveness of the safety profile.

#### 4. Exclusion Criteria

- **Weakness**: The study excluded patients with certain comorbidities, prior malignancies, and those currently using other antitumor therapies.

- **Impact**: These exclusions limit the study's applicability to the broader patient population, as the results may not be generalizable to patients with different baseline health conditions or those receiving other treatments.

**5. Potential for Selection Bias**

- **Weakness**: Stratification was based on ECOG performance status and LDH levels, but other potential confounding factors like smoking history or specific genetic markers were not considered.
- **Impact**: This may lead to selection bias, where the included patient population does not adequately represent the diversity of the real-world patient population, affecting the external validity of the study findings.

**6. Limited Endpoint Diversity**

- **Weakness**: The primary and secondary endpoints focus primarily on disease progression and response rates, with limited exploration of patient-centered outcomes like quality of life.
- **Impact**: This narrow focus may overlook important aspects of treatment impact, such as symptom relief and patient-reported outcomes, which are crucial for evaluating the overall benefit-risk profile of the treatment.

#### Conclusion:

These limitations highlight the need for cautious interpretation of the study results. Future studies should aim to include larger, more diverse patient populations, employ blinding where feasible, and extend follow-up durations to provide a more comprehensive evaluation of LY2510924's safety and efficacy. Additionally, integrating patient-centered outcomes could offer a more holistic view of the treatment's impact on patients' lives.

---

#### Study Limitations and Impact on Generalizability

**1. Study Design and Population**

- **Weakness**: The study is a Phase 2 trial with a relatively small sample size (90 patients in each treatment group), which may limit the statistical power to detect small differences between groups.
- **Impact**: The small sample size may lead to inconclusive results, particularly for secondary endpoints such as overall survival (OS) and objective response rate (ORR). Additionally, the specific inclusion and exclusion criteria (e.g., excluding patients with CNS metastases, serious concomitant systemic disorders, or recent myocardial infarction) may result in a study population that is not fully representative of the general population of patients with extensive-stage small cell lung carcinoma (ED-SCLC).

**2. Open-Label Design**

- **Weakness**: The open-label design of the study, where both participants and investigators are aware of the treatment allocations, could introduce biases, such as differential reporting of symptoms or subjective assessments of disease progression.
- **Impact**: This could affect the objectivity of outcomes like progression-free survival (PFS) and the assessment of adverse events, potentially skewing the study results.

#### 3. Adverse Event Reporting and Monitoring

- **Weakness**: Potential underreporting or inconsistent monitoring of adverse events (AEs) and serious adverse events (SAEs) across study sites may lead to an incomplete safety profile of the investigational drug, LY2510924.
- **Impact**: Inaccurate AE data can obscure the true safety profile of the treatment, affecting the risk-benefit assessment.

#### 4. Generalizability of Results

- **Weakness**: The study's exclusion criteria, such as excluding patients with certain comorbidities or those who have received prior treatment, may limit the applicability of the results to the broader patient population.
- **Impact**: The findings may not be generalizable to all patients with ED-SCLC, particularly those with more severe disease or additional health issues.

#### 5. Follow-Up and Data Collection

- **Weakness**: The follow-up period for assessing long-term outcomes, such as OS and late-onset adverse effects, may be insufficient.
- **Impact**: This limitation can prevent a comprehensive understanding of the long-term efficacy and safety of LY2510924.

### Key Takeaways and Unanswered Questions

#### Key Takeaways:

1. **Efficacy**: The addition of LY2510924 to carboplatin/etoposide demonstrated a statistically significant improvement in PFS but did not show significant benefits in secondary endpoints like ORR, OS, or DOR.
2. **Safety**: The safety profile of LY2510924 was comparable to the standard treatment, with no unexpected safety concerns identified.

#### Unanswered Questions:

1. **Long-Term Efficacy and Safety**: Further studies are needed to determine the long-term efficacy and safety of LY2510924, particularly regarding OS and potential late-onset toxicities.
2. **Biomarker Analysis**: Additional analysis is required to identify biomarkers that could predict response to LY2510924, aiding in the personalization of treatment for patients with ED-SCLC.
3. **Generalizability**: More extensive trials with broader inclusion criteria are necessary to validate the findings and ensure they are applicable to the general population of patients with ED-SCLC.

The study provides promising preliminary evidence for the use of LY2510924 in combination with carboplatin/etoposide but highlights the need for further investigation to confirm these findings and explore additional benefits.

---

### Chain of Thought Prompts

1. Define the study objective:

- What's the primary research question?
- What are the secondary outcomes of interest?

2. Review study design:
   - Is it randomized? Blinded? Controlled?
   - What's the sample size and population characteristics?
3. Examine data quality:
   - Are there missing data points?
   - How were outliers handled?
4. Analyze baseline characteristics:
   - Are treatment groups comparable?
   - Any significant differences that could impact results?
5. Evaluate primary outcome:
   - What's the main effect size?
   - Is it statistically significant?
   - What's the confidence interval?
6. Assess secondary outcomes:
   - Do they support the primary outcome?
   - Any unexpected findings?
7. Consider subgroup analyses:
   - Are effects consistent across subgroups?
   - Any notable differences?
8. Examine safety data:
   - What adverse events occurred?
   - Any serious safety concerns?
9. Analyze dropout rates:
   - How many participants completed the study?
   - Reasons for dropouts?
10. Consider potential biases:
    - Any factors that could skew results?
    - How might these impact interpretation?
11. Evaluate statistical methods:
    - Were appropriate tests used?
    - How were multiple comparisons handled?
12. Compare to similar studies:
    - How do results align with previous research?
    - Any contradictions or new insights?
13. Assess clinical significance:
    - Beyond statistical significance, what's the practical impact?
    - How might this affect patient care?

14. Consider limitations:
    - What are the study's weaknesses?
    - How might these affect generalizability?
15. Formulate conclusions:
    - What are the key takeaways?
    - What questions remain unanswered?
