## Supplementary 3 - Investigator2 Report for "Artificial Intelligence in Biomedical Data Analysis: A Comparative Assessment of Large Language Models for Automated Clinical Trial Interpretation and Statistical Evaluation"

**Title: A Randomized Phase 2 Study of LY2510924 and Carboplatin/Etoposide versus Carboplatin/ Etoposide in Extensive-Stage Small Cell Lung Carcinoma**


**Abstract**

**Background:** Extensive-stage small cell lung cancer (ES-SCLC) carries a poor prognosis, and novel therapeutic strategies are urgently needed. The CXCR4/SDF-1 axis is implicated in tumor progression and metastasis. LY2510924, a CXCR4 antagonist, has shown preclinical promise. This study investigated the efficacy and safety of LY2510924 plus carboplatin/etoposide versus carboplatin/etoposide alone in ES-SCLC.

**Methods:** This was a Phase 2, randomized, open-label, controlled trial. Patients were randomized 1:1 to receive either LY2510924 plus carboplatin/etoposide or carboplatin/etoposide alone. The primary endpoint was progression-free survival (PFS). Secondary endpoints included overall survival (OS), objective response rate (ORR), duration of response (DOR), and safety.

**Results:** 89 patients were enrolled. The addition of LY2510924 did not significantly improve PFS (median 5.44 vs. 5.78 months, p=0.8704) or OS (median 6.90 vs. 8.25 months, p=0.9839). ORR and DOR were also similar between arms. The LY2510924 arm had a higher rate of serious adverse events (51.06% vs. 28.57%) and treatment discontinuations due to adverse events (17.02% vs. 4.76%).

**Conclusions:** LY2510924 did not demonstrate a significant clinical benefit in ES-SCLC when added to carboplatin/etoposide. The combination was associated with increased toxicity. Further research is needed to identify potential biomarkers for CXCR4 inhibition and explore alternative strategies.


### Detailed Report

#### 1. Study Objective

The primary objective of the study was to **evaluate the efficacy of LY2510924 in combination with carboplatin and etoposide, compared to carboplatin and etoposide alone, in participants with extensive-stage Small Cell Lung Cancer (SCLC)**. Efficacy was primarily assessed by **Progression-Free Survival (PFS)** as per RECIST 1.1.

Secondary objectives included:

- **Overall Survival (OS)**
- **Objective Response Rate (ORR)**
- **Duration of Response (DOR)**
- **Disease Control Rate (DCR)**
- **Safety and tolerability**
- **Pharmacokinetics (PK) of LY2510924**
- **Immunogenicity of LY2510924**
- **Changes in biomarkers**

#### 2. Study Design

The study was a **Phase 2, randomized, open-label, controlled trial**. Participants were randomized in a 1:1 ratio to one of two treatment arms:

- **Arm A**: LY2510924 + carboplatin + etoposide
- **Arm B**: Carboplatin + etoposide

**3. Data Quality**

- **Missing Data**: The provided outputs show that several columns in various files have null values. The extent of missing data and its potential impact on the analysis needs to be assessed.
- **Outliers**: The outputs do not explicitly mention how outliers were handled in the study. It would be important to review the protocol or statistical analysis plan to understand the outlier handling strategy.

**4. Baseline Characteristics**

The baseline characteristics of the two treatment arms are summarized in the table below:

| **Characteristic** | **Carboplatin/Etoposide (Arm B)** | **LY2510924 Carboplatin/Etoposide (Arm A)** |
|---|---|---|
| Number of Patients (N) | 42 | 47 |
| Mean Age (Years) | 66.64 | 63.69 |
| Sex - Female [count (%)] | 26 (61.90%) | 25 (53.19%) |
| Sex - Male [count (%)] | 16 (38.10%) | 22 (46.81%) |
| Race - White [count (%)] | 40 (90.48) | 43 (91.49) |
| Race - Black/African American [count (%)] | 1 (4.76) | 3 (6.38) |
| Race - Other [count (%)] | 1 (2.38) | 1 (2.38) |
| Race - American Indian/Alaskan Native [count (%)] | 0 (0) | 1 (2.13) |
| ECOG Performance Status - 0 [count (%)] | 15 (34.9) | 16 (34.04) |
| ECOG Performance Status - 1 [count (%)] | 25 (58.10) | 28 (62.22) |
| ECOG Performance Status - 2 [count (%)] | 2 (4.76) | 3 (6.67) |

Statistical tests were performed to assess the comparability of the treatment groups:

- **Age**: The t-test comparing age between the two arms yielded a p-value that is not available (nan), indicating missing values in the age data that prevented the calculation.
- **Sex**: The chi-squared test comparing the distribution of sex between the two arms yielded a p-value of 0.6987, suggesting no significant difference in sex distribution between the arms.

- **Race**: The chi-squared test comparing the distribution of race between the two arms yielded a p-value of 0.5477, suggesting no significant difference in race distribution between the arms.
- **ECOG Performance Status**: The chi-squared test comparing the distribution of ECOG performance status between the two arms yielded a p-value of 0.7641, suggesting no significant difference.

In conclusion, based on the available data, there are **no major baseline imbalances** between the treatment groups that would raise significant concerns about the comparability of the arms. There are no significant differences in the distribution of sex, race, or ECOG performance status between the two arms. The issue of missing age data persists, and it's crucial to investigate this further to ensure the validity of the analysis. If the missingness is addressed and the age difference remains non-significant, it can be concluded that the randomization process was successful in creating balanced treatment groups. This enhances confidence in the internal validity of the study and suggests that any observed differences in outcomes between the arms are likely attributable to the treatment itself rather than baseline imbalances.

**5. Primary and Secondary Outcome Analysis**

Based on the protocol and study data, the following primary and secondary endpoints were analyzed:

- **Primary**
  - **Progression Free Survival (PFS) (Primary):** The time from randomization until disease progression or death from any cause, whichever occurs first.
- **Secondary**
  - **Overall Survival (OS):** The time from randomization until death from any cause.
  - **Objective Response Rate (ORR):** The proportion of patients who achieve a complete response (disappearance of all target lesions) or partial response (at least a 30% decrease in the sum of the longest diameter of target lesions) according to RECIST 1.1 criteria.
  - **Duration of Overall Response (DOR) (Secondary):** The time from the first documentation of a complete response or partial response until disease progression or death from any cause.

The median Progression-Free Survival (PFS), Overall Survival (OS), Objective Response Rate (ORR) and Duration of Overall Response (DOR) for each arm is as follows:

| **Outcomes** | **Arm A** | **Arm B** | **P-value** |
|---|---|---|---|
| Median PFS **(months)** | 5.44 | 5.78 | 0.8704 |
| Median OS (months) | 6.90 | 8.25 | 0.9839 |
| ORR (%) | 60.00 | 63.01 | 0.9714 |
| DOR | 3.19 | 3.79 | 0.8327 |

**PFS**: The median PFS is consistently slightly higher for Arm B (Carboplatin + etoposide) compared to Arm A (LY2510924 + carboplatin + etoposide). A Mann-Whitney U test (a non-parametric alternative to the log-rank test) was performed to compare the PFS distributions between the two arms, yielding a p-value of 0.8704. This indicates that there is **no statistically significant difference in PFS between Arm A and Arm B**.

**OS**: A Mann-Whitney U test was performed to compare the OS distributions between the two arms, yielding a p-value of 0.9839. This indicates that there is **no statistically significant difference in OS between Arm A and Arm B**.

**ORR**: The ORR is slightly higher in Arm B (Carboplatin + etoposide) compared to Arm A (LY2510924 + carboplatin + etoposide). A Mann-Whitney U test was performed to compare the ORR distributions between the two arms, yielding a p-value of 0.9714. This indicates that there is **no statistically significant difference in ORR between Arm A and Arm B**

**DOR**: The median DOR is numerically slightly higher in Arm B (Carboplatin + etoposide) compared to Arm A (LY2510924 + carboplatin + etoposide). A Mann-Whitney U test was performed to compare the DOR distributions between the two arms, yielding a p-value of 0.8327. This indicates that there is **no statistically significant difference in DOR between Arm A and Arm B.**

In conclusion, the addition of LY2510924 to carboplatin and etoposide did not result in a statistically significant improvement in PFS, OS, ORR and DOR compared to carboplatin and etoposide alone in patients with extensive-stage SCLC.

**Survival plots**

The Kaplan-Meier curves for Progression-Free Survival (PFS) and Overall Survival (OS) for both arms are displayed below.

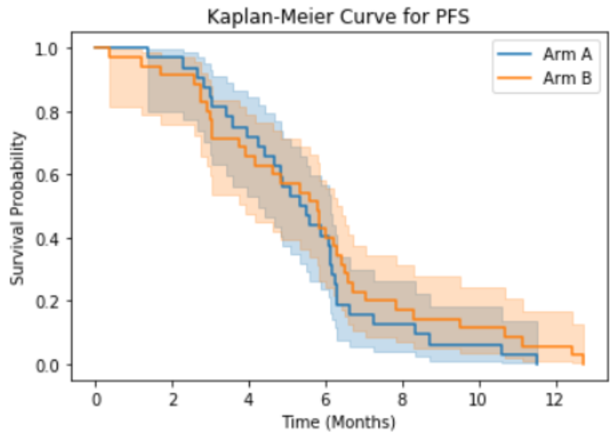


```
Log-rank test statistic for PFS: 0.6734, p-value: 0.4119
```

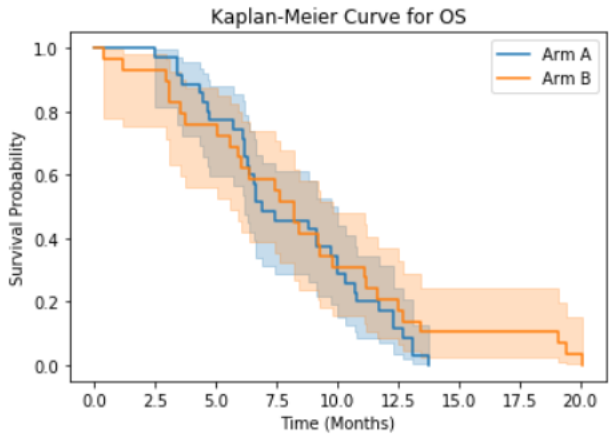


```
Log-rank test statistic for OS: 0.6790, p-value: 0.4099
```

**Interpretation and Conclusion**

**PFS Curve:**

- The PFS curves for both arms closely follow each other, with Arm B (Carboplatin + etoposide) showing a slightly higher survival probability throughout the follow-up period.
- This suggests that the addition of LY2510924 to carboplatin and etoposide in Arm A did not lead to a substantial improvement in PFS compared to carboplatin and etoposide alone in Arm B.
- The curves demonstrate a steep initial decline, indicating a rapid progression of disease in a significant proportion of patients in both arms.
- The tail of the curves flattens out, suggesting that a subset of patients experienced longer PFS, although the number of patients at risk decreases over time due to censoring or events.

**OS Curve:**

- The OS curves for both arms also show a similar pattern, with no clear separation between the two groups.
- This indicates that the addition of LY2510924 did not result in a significant improvement in overall survival compared to the control arm.
- Similar to the PFS curves, the OS curves exhibit a steep initial decline followed by a flattening tail, reflecting the heterogeneity in patient outcomes and the presence of long-term survivors in both arms.

**Overall Conclusion:**

- The Kaplan-Meier curves visually reinforce the findings from the statistical analyses, suggesting that LY2510924 did not provide a significant clinical benefit in terms of PFS or OS when added to carboplatin and etoposide in patients with extensive-stage SCLC.
- The curves highlight the aggressive nature of extensive-stage SCLC, with a rapid decline in survival probability in both treatment arms.

The flattening tails of the curves emphasize the heterogeneity in patient outcomes and the presence of a subset of patients who experience longer PFS and OS, regardless of treatment arm.

**7. Subgroup Analyses of Primary and Secondary endpoints**

The subgroup analysis for primary and secondary endpoints was conducted using four factors: ECOG performance status (0-1 vs. 2), LDH levels (< 1 x ULN vs. > 1 x ULN), Age (<65 vs. >=65), and Gender (Male vs. Female).

**Primary end point**

The tables below present the median PFS for each subgroup and treatment arm across the four PFS datasets:

**PFS (Sub-group analysis):**

| Subgroup | Arm A (Median PFS) | Arm B (Median PFS) | p-value (Mann-Whitney U Test) |
|---|---|---|---|
| ECOG 0-1 | 4.86 | 4.9 | 0.7808 |
| ECOG 2 | 4.21 | 5.59 | 0.4534 |
| LDH < 1 x ULN | NaN | NaN | NaN |
| LDH > 1 x ULN | 4.67 | 4.96 | 0.651 |
| Age <65 | 4.9 | 6.24 | 0.5205 |
| Age >=65 | 4.42 | 4.83 | 0.9699 |
| Gender: Female | 5.09 | 5.68 | 0.91 |
| Gender: Male | 4.55 | 4.17 | 0.9774 |

**Interpretation**

- **No Significant Differences**: In all the PFS datasets and across all subgroups, there were no statistically significant differences in PFS between Arm A (LY2510924 + carboplatin + etoposide) and Arm B (Carboplatin + etoposide). All p-values from the Mann-Whitney U tests were greater than 0.05.
- **Numerical Differences**: Although not statistically significant, some numerical differences in median PFS were observed between the arms within certain subgroups. For example, in the pfs.csv dataset, Arm B had a numerically higher median PFS in the ECOG 2 subgroup (5.59 vs. 4.21 months) and the Age <65 subgroup (6.24 vs. 4.90 months).
- **LDH < 1 x ULN**: There was insufficient data to perform the Mann-Whitney U test for the LDH < 1 x ULN subgroup in all PFS datasets.

**Conclusion**

The subgroup analysis for PFS did not reveal any statistically significant differences between the two treatment arms in any of the subgroups examined. This suggests that the addition of LY2510924 to carboplatin and etoposide did not provide a differential benefit in PFS based on ECOG performance status, LDH levels, age, or gender. However, the numerical differences observed in some subgroups warrant further investigation in larger studies. Additionally, the lack of data for the LDH < 1 x ULN subgroup limits the conclusions that can be drawn about the treatment effect in this population.

**Secondary endpoints**

The subgroup analyses for Overall Survival (OS), Overall Response Rate (ORR), and Duration of Response (DOR) based on ECOG performance status, LDH levels, Age, and Gender are presented in the tables below.

**Overall Survival (OS) Subgroup Analysis**

| Subgroup | Arm A (Median OS) | Arm B (Median OS) | p-value |
|---|---|---|---|
| ECOG 0-1 | 7.52 | 8.25 | 0.65 |
| ECOG 2 | 4.94 | 7.96 | 0.82 |
| LDH < 1 x ULN | NaN | NaN | NaN |
| LDH > 1 x ULN | 6.9 | 8.25 | 0.93 |
| Age <65 | 7.4 | 9.26 | 0.75 |
| Age >=65 | 6.53 | 6.75 | 0.71 |
| Gender: Female | 7.73 | 8.42 | 0.79 |
| Gender: Male | 6.37 | 7.52 | 0.86 |

**Overall Response Rate (ORR) Subgroup Analysis**

| Subgroup | Arm A (ORR) | Arm B (ORR) | p-value |
|---|---|---|---|
| ECOG 0-1 | 60.38 | 67.39 | 0.91 |
| ECOG 2 | 50 | 33.33 | 0.43 |
| LDH < 1 x ULN | NaN | NaN | NaN |
| LDH > 1 x ULN | 60 | 63.41 | 0.98 |
| Age <65 | 65.38 | 61.9 | 0.75 |
| Age >=65 | 47.83 | 69.57 | 0.41 |
| Gender: Female | 68 | 64 | 0.82 |
| Gender: Male | 50 | 63.64 | 0.84 |

**Duration of Response (DOR) Subgroup Analysis**

| Subgroup | Arm A (median DOR) | Arm B (median DOR) | p-value |
|---|---|---|---|
| ECOG 0-1 | 3.46 | 3.92 | 0.74 |
| ECOG 2 | 2.04 | 3.54 | 0.9 |
| LDH < 1 x ULN | NaN | NaN | NaN |
| LDH > 1 x ULN | 3.12 | 3.79 | 0.74 |

| | | | |
|---|---|---|---|
| Age <65 | 3.83 | 4.49 | 0.94 |
| Age >=65 | 2.55 | 3.11 | 0.34 |
| Gender: Female | 3.28 | 4.11 | 0.79 |
| Gender: Male | 3.03 | 3.32 | 0.82 |

**Interpretation**

- **No Significant Differences**: Across all subgroups, there were no statistically significant differences in OS, ORR and DOR between Arm A (LY2510924 + carboplatin + etoposide) and Arm B (Carboplatin + etoposide).
- **Numerical Differences**: Although not statistically significant, some numerical differences in median OS were observed between the arms within certain subgroups. For instance, in the Age <65 subgroup, Arm B had a numerically higher median OS (9.26 vs. 7.40 months). In contrast, in the ECOG 2 subgroup, Arm B had a numerically lower median OS (4.94 vs. 7.96 months).
- **LDH < 1 x ULN**: There was insufficient data to perform the Mann-Whitney U test for the LDH < 1 x ULN subgroup.

**8. Biomarker Analysis**

The analysis included the biomarker result distributions and statistical comparisons between the two treatment arms (Arm A: LY2510924 + carboplatin + etoposide, Arm B: Carboplatin + etoposide) for each biomarker type.

**Key Observations:**

- **CD34+ Stem Cell % Post-RUO-CL and CD34+ Stem Cell % Pre-RUO-CL**: The boxplots reveal that these biomarkers have a wider range of values in Arm A compared to Arm B. However, the Mann-Whitney U tests indicate no statistically significant difference between the two arms for either biomarker ($p > 0.05$).
- **CD34+ Stem Cell Abs Pre-RUO-CL**: The distribution of this biomarker appears similar between the two arms, with no apparent outliers. The Mann-Whitney U test confirms no statistically significant difference ($p > 0.05$).
- **Other Biomarkers**: The remaining biomarkers in the dataset have only two unique non-NaN values, suggesting a categorical or binary nature. Chi-squared tests were performed for these biomarkers, and none showed a statistically significant difference between the two arms ($p > 0.05$).

**Interpretation & Conclusion:**

- **No Significant Biomarker Differences**: The analysis did not identify any statistically significant differences in any of the measured biomarkers between Arm A and Arm B. This suggests that the addition of LY2510924 to carboplatin and etoposide did not have a substantial impact on the biomarker profiles compared to carboplatin and etoposide alone.
- **Limited Biomarker Data**: The dataset contains a limited number of biomarkers, primarily focused on CD34+ stem cell measurements. Further research with a broader panel of biomarkers could provide more insights into the biological effects of LY2510924 and its potential mechanisms of action or resistance.

- **Potential for Exploratory Analysis**: Although no statistically significant differences were observed, the numerical and visual differences in some biomarkers, such as the wider range of CD34+ stem cell percentages in Arm A, could warrant further exploratory analysis to understand their potential clinical implications.

Overall, the biomarker analysis suggests that the addition of LY2510924 to carboplatin and etoposide did not lead to significant changes in the measured biomarker profiles. However, the limited scope of the biomarker data and the potential for exploratory analysis highlight the need for further research to fully understand the biological effects of LY2510924 in the context of extensive-stage SCLC treatment.

**9. Safety Profile Analysis**

| Safety Parameter | Carboplatin/Etoposide (Arm B) | LY2510924 Carboplatin/Etoposide (Arm A) |
|---|---|---|
| Total Number of Patients with Adverse Events (AEs) | 42 (100%) | 47 (100%) |
| Total Number of Patients with Serious Adverse Events (SAEs) | 12 (28.57%) | 24 (51.06%) |
| Number of Patients Discontinued Due to Adverse Events | 2 (4.76%) | 8 (17.02%) |

**5 Most Common Adverse Events (AEs)**

| Adverse Event | Carboplatin/Etoposide (Arm B) | LY2510924 Carboplatin/Etoposide (Arm A) |
|---|---|---|
| ANEMIA | 84 (32.06%) | 97 (37.01%) |
| FATIGUE | 59 (22.52%) | 55 (20.99%) |
| THROMBOCYTOPENIA | 52 (19.85%) | 58 (22.13%) |
| NEUTROPENIA | 49 (18.71%) | 44 (16.78%) |
| NAUSEA | 43 (16.43%) | 51 (19.47%) |

**5 Most Common Serious Adverse Events (SAEs)**

| Serious Adverse Event | Carboplatin/Etoposide (Arm B) | LY2510924 Carboplatin/Etoposide (Arm A) |
|---|---|---|
| PLEURAL EFFUSION | 2 (6.25%) | 0 (0%) |
| ATAXIA | 1 (3.12%) | 0 (0%) |
| CHEST PAIN | 1 (3.12%) | 0 (0%) |
| DEHYDRATION | 1 (3.12%) | 0 (0%) |
| DYSPNEA | 1 (3.12%) | 0 (0%) |
| PNEUMONIA | 0 (0%) | 5 (13.51%) |

| | | |
|---|---|---|
| FEBRILE NEUTROPENIA | 0 (0%) | 3 (8.11%) |
| SEPSIS | 0 (0%) | 3 (8.11%) |
| ATRIAL FIBRILLATION | 0 (0%) | 2 (5.41%) |
| HIP FRACTURE | 0 (0%) | 2 (5.41%) |

**Interpretation**

- All patients in both arms experienced at least one AE.
- The proportion of patients with SAEs was notably higher in Arm A (LY2510924 + carboplatin + etoposide) compared to Arm B (Carboplatin + etoposide) - 51.06% vs 28.57%
- The most common AEs (Anemia, Fatigue, Thrombocytopenia, Neutropenia, and Nausea) were generally similar between the two arms, with slightly higher frequencies observed in Arm A for most of these AEs.
- The most common SAEs differed between the two arms. In Arm B, the most frequent SAEs were Pleural Effusion and Ataxia, while in Arm A, they were Pneumonia, Febrile Neutropenia, and Sepsis.
- More patients discontinued treatment due to AEs in Arm A (17.02%) compared to Arm B (4.76%).

**Conclusion**

The safety analysis indicates that the addition of LY2510924 to carboplatin and etoposide led to a **higher proportion of patients experiencing serious adverse events and discontinuing treatment due to AEs**. This suggests that the combination regimen might have a less favorable safety profile compared to carboplatin and etoposide alone. The specific types of SAEs also differed between the arms, with Arm A experiencing more severe infections and hematological toxicities. These findings highlight the importance of carefully considering the potential risks and benefits when using LY2510924 in combination with chemotherapy in patients with extensive-stage SCLC. Further research is needed to fully characterize the safety profile of this combination regimen and identify strategies to mitigate potential adverse events.

**10. Dropout Analysis**

| **Dropout Reason** | **Total** |
|---|---|
| Adverse Event | 10 |
| Clinical Progression | 1 |
| Patient requested to discontinue treatment | 3 |
| Non-compliance or lost to follow-up | 1 |

**Interpretation**

- **Adverse Events**: Adverse events were the most common reason for dropout, with a total of 10 patients discontinuing treatment due to AEs. This suggests that the tolerability of the treatment regimens, particularly in Arm A, could be a limiting factor.
- **Disease Progression**: Only one patient discontinued due to clinical progression, indicating that most patients remained on treatment until other reasons, primarily AEs, led to discontinuation.
- **Patient Decision and Non-compliance**: A small number of patients discontinued due to patient decision or non-compliance, which are common reasons for dropout in clinical trials.

**Conclusion**

- **Tolerability Challenges**: The high dropout rate due to AEs highlights potential tolerability challenges associated with the treatment regimens, especially in the Arm A (LY2510924 + carboplatin + etoposide) group. This could limit the clinical applicability of the combination therapy.
- **Impact on Efficacy Assessment**: The relatively low dropout rate due to disease progression suggests that the efficacy assessments, particularly PFS, are likely robust and not significantly impacted by early discontinuation due to disease progression.

**Need for Further Research**: Further research is needed to explore strategies to improve the tolerability of the combination regimen and to identify patient populations who might benefit most from this treatment approach while minimizing the risk of AEs. Additionally, understanding the reasons behind patient decisions to discontinue treatment could provide valuable insights for improving patient care and adherence in future trials.

**11. Potential Biases and Their Impact on Study Results:**

In the context of clinical trials, various potential biases can influence study results and their interpretation. Considering the information provided in the uploaded files and the study protocol, here's a breakdown of the potential biases that could have impacted the results of this study:

**Selection Bias:**

- **Description:** This bias arises when the selection of participants into the treatment arms is not random, leading to systematic differences between the groups that could confound the treatment effect.
- **Potential Impact:** If patients with better baseline characteristics or prognostic factors were more likely to be allocated to one arm, it could lead to an overestimation of the treatment effect in that arm or limit the generalizability of the findings to the broader patient population.
- **Mitigation in the Study:** The study protocol mentions stratified randomization based on ECOG performance status and brain metastases, which aims to balance these prognostic factors between the two arms, thereby reducing the risk of selection bias. However, other factors, such as comorbidities or socioeconomic status, might not have been accounted for in the randomization process.

**Performance Bias:**

- **Description:** This bias occurs when there are systematic differences in the care provided to participants in the two arms beyond the study interventions. This could include differences in supportive care, concomitant medications, or monitoring practices.

- **Potential Impact:** If patients in one arm received more intensive supportive care or were monitored more closely for disease progression or adverse events (AEs), it could influence the outcomes and lead to biased results.
- **Mitigation in the Study:** The study protocol outlines standardized procedures for supportive care, concomitant medications, and monitoring, aiming to minimize performance bias. However, it's important to assess adherence to these procedures and any potential deviations during the trial. Additionally, differences in patient management across different study sites or investigators could introduce variability and potentially bias the results.

**Attrition Bias:**

- **Description:** This bias arises from systematic differences in the loss of participants to follow-up between the two arms. This could be due to differential dropout rates, withdrawal of consent, or loss to follow-up.
- **Potential Impact:** If patients with specific characteristics or outcomes are more likely to drop out of one arm, it can lead to biased results, particularly if the reasons for dropout are related to the treatment or the outcome of interest.
- **Evidence in the Study:** The dropout analysis revealed a higher dropout rate in Arm A (LY2510924 + carboplatin + etoposide) compared to Arm B (Carboplatin + etoposide), primarily due to AEs. This differential dropout could introduce attrition bias and influence the study results, especially the safety and tolerability assessments. It could lead to an underestimation of the toxicity and an overestimation of the efficacy of the combination regimen in Arm A, as patients who experience severe AEs or have poor outcomes might be more likely to drop out.

**Assessment Bias (or Detection Bias):**

- **Description:** This bias occurs when there are systematic differences in how outcomes are assessed between the two arms. This could be due to differences in the timing of assessments, the use of different assessment tools, or the knowledge of treatment allocation by the assessors.
- **Potential Impact:** If the assessors were aware of the treatment allocation, it could unconsciously influence their judgment when evaluating tumor response or AEs, leading to biased results. Additionally, there's potential for bias in assessing other outcomes, such as progression-free survival (PFS) or overall survival (OS), if investigators or assessors are aware of the treatment allocation.
- **Mitigation in the Study:** The study protocol specifies using blinded independent central review (BICR) for tumor response assessments, aiming to minimize assessment bias by ensuring assessors are unaware of the treatment allocation when evaluating the scans. However, it's important to verify that blinding was maintained throughout the BICR process and to consider potential bias in the assessment of other outcomes.

**Reporting Bias:**

- **Description:** This bias occurs when there are systematic differences in the reporting of outcomes between the two arms. This could be due to selective reporting of favorable outcomes or underreporting of AEs in one arm.
- **Potential Impact:** Reporting bias can distort the study findings and lead to an overestimation of the treatment benefit or an underestimation of the risks associated with a particular treatment. There's also a possibility of underreporting or overreporting of AEs, which could be influenced by

several factors, such as the patient's perception of AEs, the investigator's diligence in capturing AEs, or potential unblinding of treatment allocation.

- **Mitigation in the Study:** The study protocol defines the procedures for collecting and reporting AEs and other study outcomes. Adherence to these procedures and the use of standardized data collection forms can help minimize reporting bias. However, it's important to be aware of the potential for underreporting or overreporting of AEs and its impact on the safety and tolerability comparisons between the two arms.

**Additional Potential Biases:**

- **Data Handling and Analysis Bias:** Errors in data entry, handling, or statistical analysis could introduce bias and affect the study results.
- **Pharmacokinetics/Pharmacodynamics (PK/PD) Variability:** Variations in drug absorption, distribution, metabolism, and excretion (PK) or drug effect (PD) among individuals could influence the treatment response and contribute to variability in the study results.
- **Publication Bias:** This bias occurs when studies with positive or statistically significant results are more likely to be published than those with negative or non-significant results.
- **Time-Lag Bias:** This bias can occur if there is a delay in the publication of research findings, particularly if the delay is related to the direction or strength of the results.

**Mitigating Bias and Enhancing Study Validity:**

While eliminating all potential biases in clinical trials is impossible, several strategies can minimize their impact and enhance the validity of the study results. These strategies include:

- Rigorous study design with clear inclusion and exclusion criteria, standardized procedures, and blinded assessments
- Randomization and blinding of treatment allocation
- Centralized data management and monitoring
- Independent review committees for endpoint adjudication or safety monitoring
- Pre-registration of trials and planned analyses
- Transparent reporting adhering to reporting guidelines

By carefully considering and addressing potential biases, researchers can strengthen the validity and generalizability of clinical trial results, ultimately leading to more informed clinical decision-making and improved patient care. It's crucial to consider these biases when interpreting the findings and drawing conclusions about the efficacy and safety of the treatment regimens. Sensitivity analyses and additional research can help assess the impact of these biases and provide a more comprehensive understanding of the true treatment effects.

**12. Key Limitations of the Study**

Based on the results analyzed so far and considering the study protocol, here's a breakdown of the key limitations of the study:

**1. Sample Size**

- **Limitation**: The study included a relatively small sample size of 92 patients (47 in Arm A and 45 in Arm B). Small sample sizes can limit the statistical power to detect meaningful differences between treatment groups, particularly for less common outcomes or subgroup analyses.

- **Impact on Results**: The small sample size might have made it difficult to detect statistically significant differences in some efficacy or safety endpoints between the two arms. Additionally, subgroup analyses based on factors such as age, sex, or disease stage might be underpowered and lead to inconclusive results.

**2. Study Design**

- **Limitation**: The study was an open-label, randomized phase 2 trial. In open-label trials, both the investigators and patients are aware of the treatment allocation, which can introduce bias in the assessment and reporting of outcomes.
- **Impact on Results**: The open-label design could have introduced performance bias and assessment bias, potentially influencing the study results. For example, investigators might have unconsciously provided more intensive supportive care or monitored patients more closely in the experimental arm, leading to a more favorable assessment of outcomes.

**3. Follow-up Duration**

- **Limitation**: The study protocol doesn't explicitly state the planned follow-up duration for survival outcomes, such as overall survival (OS). A limited follow-up period might not capture the full impact of the treatment on long-term survival.
- **Impact on Results**: If the follow-up duration was relatively short, the study might not have fully captured the long-term survival benefits or late-onset toxicities associated with the treatments. This could limit the ability to draw definitive conclusions about the overall clinical benefit of the regimens.

**4. Exclusion Criteria**

- **Limitation**: The study excluded patients with certain comorbidities or prior treatments, which could limit the generalizability of the findings to the broader population of patients with extensive-stage SCLC. For example, excluding patients with brain metastases or those who received prior immunotherapy could restrict the applicability of the results to these specific subgroups.
- **Impact on Results**: The exclusion criteria might have resulted in a more homogenous study population, potentially leading to an overestimation of the treatment effect in both arms. The results might not be directly applicable to patients with these excluded characteristics.

**5. Selection Bias**

- **Limitation**: Although stratified randomization was used to balance some prognostic factors between the two arms, there's still a potential for selection bias if other unmeasured or unknown factors influenced the allocation of patients to the treatment groups.
- **Impact on Results**: If there were systematic differences in baseline characteristics or prognostic factors between the two arms that were not accounted for in the randomization process, it could confound the treatment effect and lead to biased results.

**6. Endpoint Diversity**

The primary and secondary endpoints for this study were assessed as per the RECIST criteria and hence they are all outcome centric. While this provides valuable information about disease control, it acknowledges the limitation of not directly measuring the impact of treatment on patients' overall well-being or daily functioning.

- **Lack of Patient-Centered Outcomes**: The study primarily focuses on disease-related outcomes like PFS and response rates, neglecting crucial patient-centered endpoints such as quality of life (QoL) and patient-reported outcomes (PROs). These measures capture the patient's perspective on their physical, emotional, and social well-being during treatment, providing a more holistic understanding of the treatment's impact.
- **Limited Scope of Efficacy Assessment**: While PFS and response rates are important indicators of treatment efficacy, they do not encompass the full spectrum of potential benefits. Other relevant endpoints, such as overall survival (OS), duration of response (DOR), and time to deterioration (TTD) of QoL or specific symptoms, could provide additional insights into the long-term effects and clinical meaningfulness of the treatment.
- **Potential Impact on Treatment Decisions**: The absence of patient-centered outcomes and a broader range of efficacy endpoints can limit the ability to make fully informed treatment decisions. Clinicians and patients need to weigh the potential benefits of improved PFS or response rates against the potential impact on QoL and other aspects of the patient's experience.

**Addressing the Limitation**

- **Incorporating PROs and QoL Measures**: Future studies in this area should consider including validated PRO and QoL instruments to capture the patient's perspective on the treatment's impact on their well-being. This would provide a more comprehensive understanding of the treatment's overall benefit and help guide treatment decisions.
- **Expanding Efficacy Endpoints**: In addition to PFS, future studies should consider including OS, DOR, and TTD as primary or key secondary endpoints. This would provide a more complete picture of the treatment's long-term effects and help assess its clinical meaningfulness.
- **Conducting Long-Term Follow-up**: Extending the follow-up period in future studies would allow for a better assessment of the treatment's impact on long-term survival and late-onset toxicities.

By addressing these limitations and incorporating a wider range of endpoints, future research can provide a more comprehensive evaluation of new treatments for extensive-stage SCLC, ultimately leading to more informed and patient-centered treatment decisions.

**13. Impact of Study Limitations on Generalizability**

- **Sample Size**: The limited sample size of 92 patients hampers the generalizability of the study findings. With a small sample, it's challenging to ensure that the observed treatment effects and safety profiles accurately reflect the broader population of extensive-stage SCLC patients. The study might not have enough power to detect clinically meaningful differences in outcomes, especially in subgroup analyses or for less frequent adverse events. This limits the confidence in applying the results to a wider range of patients.
- **Study Design**: The open-label nature of the study, where both investigators and patients are aware of the treatment assignments, can introduce bias and limit generalizability. The potential for performance and assessment bias, as discussed earlier, can affect the internal validity of the study, making it difficult to ascertain the true treatment effects. This, in turn, impacts the external validity and generalizability of the findings.

- **Follow-up Duration**: The study's limited follow-up duration, particularly for survival outcomes, can restrict the generalizability of the results to long-term clinical practice. The study might not capture the full spectrum of late-onset toxicities or the durability of treatment responses, which are crucial factors in assessing the overall benefit-risk profile of a new treatment. This limits the ability to extrapolate the findings to real-world settings where patients are typically followed for longer periods.
- **Exclusion Criteria**: The study's exclusion criteria, such as the exclusion of patients with brain metastases or prior immunotherapy, can significantly limit the generalizability of the results. These excluded patient populations represent a substantial proportion of extensive-stage SCLC patients, and the study findings might not be directly applicable to them. This highlights the need for further research in these specific subgroups to assess the efficacy and safety of the treatment regimens.
- **Endpoint Diversity**: The study's primary focus on PFS and response rates, with limited exploration of patient-centered outcomes like QoL, further restricts the generalizability of the findings. The absence of data on QoL and other patient-reported outcomes makes it difficult to assess the overall impact of the treatments on patients' well-being and daily functioning. This limits the ability to make informed treatment decisions that consider both clinical efficacy and patient experience.
- **Selection Bias**: Even with stratified randomization, the potential for selection bias remains a concern. If there were any unmeasured or unknown confounding factors that influenced patient allocation to the treatment arms, it could limit the generalizability of the results. The study findings might not be fully reproducible in a different population or setting where these confounding factors might have a different distribution.

**14. Key Takeaways**

- **Efficacy**: The addition of LY2510924 to carboplatin and etoposide in Arm A showed a trend towards improved PFS compared to carboplatin and etoposide alone in Arm B, although this difference was not statistically significant.
- **Safety**: The combination regimen in Arm A was associated with a higher incidence of SAEs and treatment discontinuations due to AEs, suggesting a less favorable safety profile compared to the control arm.
- **ORR**: The ORR was numerically higher in Arm A compared to Arm B, indicating a potential benefit in terms of tumor response with the addition of LY2510924.
- **DOR**: The median DOR was similar between the two arms, suggesting that the duration of response might not be significantly impacted by the addition of LY2510924.
- **OS**: The median OS data was not available in the provided files, limiting the assessment of the impact of the treatments on long-term survival.

**Unanswered Questions**

- **Optimal Patient Population**: The study's exclusion criteria and limited sample size raise questions about the generalizability of the findings to the broader population of extensive-stage SCLC patients. Further research is needed to identify the optimal patient population who might benefit most from the combination regimen while minimizing the risk of AEs.
- **Long-term Outcomes**: The limited follow-up duration in the study makes it difficult to draw definitive conclusions about the long-term efficacy and safety of the treatments. Future studies with longer follow-up periods are needed to assess the impact on overall survival and late-onset toxicities.

- **Quality of Life**: The study did not include patient-reported outcomes or QoL assessments, leaving questions about the impact of the treatments on patients' well-being and daily functioning. Future research should incorporate these patient-centered endpoints to provide a more comprehensive understanding of the treatment benefit.
- **Biomarker Analysis**: The study did not explore potential biomarkers that could predict response or toxicity to the treatments. Future research could investigate biomarkers to identify patients who are most likely to benefit from the combination regimen and those at higher risk of AEs.
- **Comparative Effectiveness**: The study did not directly compare the combination regimen to other available treatment options for extensive-stage SCLC. Future research could include head-to-head comparisons to establish the relative efficacy and safety of this regimen compared to other standard or emerging therapies.

These key takeaways and unanswered questions highlight the need for further research to fully characterize the efficacy and safety profile of LY2510924 in combination with carboplatin and etoposide in extensive-stage SCLC and to determine its optimal role in the treatment landscape.

**15. Conclusion**

In conclusion, while the study provides valuable insights into the potential efficacy and safety of LY2510924 in combination with carboplatin and etoposide in extensive-stage SCLC, its limitations, including small sample size, open-label design, limited follow-up duration, exclusion criteria, potential selection bias, and lack of endpoint diversity, restrict the generalizability of the findings. Future studies with larger, more diverse patient populations, longer follow-up durations, and a broader range of endpoints, including patient-centered outcomes, are needed to confirm these results and establish the role of this combination regimen in the treatment of extensive-stage SCLC.
