## Supplementary 4 - Investigator3 Report for "Artificial Intelligence in Biomedical Data Analysis: A Comparative Assessment of Large Language Models for Automated Clinical Trial Interpretation and Statistical Evaluation"

### A Randomized Phase 2 Study of LY2510924 and Carboplatin/Etoposide versus Carboplatin/Etoposide in Extensive-Stage Small Cell Lung Carcinoma: An Integrated Analysis

September 23, 2024


#### Abstract

This study investigated the efficacy and safety of LY2510924, a CXCR4 antagonist, combined with carboplatin/etoposide versus carboplatin/etoposide alone in extensive-stage small-cell lung carcinoma (ED-SCLC). The primary endpoint was progression-free survival (PFS); secondary endpoints included objective response rate (ORR), overall survival (OS), and safety. The multicenter, randomized, open-label Phase 2 trial included 129 patients. The median PFS was 4.67 months in Arm A (LY2510924 + carboplatin/etoposide) versus 5.14 months in Arm B (carboplatin/etoposide alone) (p=0.56). ORR was 0% in both arms. Median OS was 9.17 months in Arm A compared to 9.25 months in Arm B (p=0.74). Common adverse events (AEs) were not captured due to data limitations in the AETERM column. However, the most frequent high-level group terms for adverse events included 'General disorders and administration site conditions' (21.4%), 'Blood and lymphatic system disorders' (14.5%), and 'Gastrointestinal disorders' (13.3%). The dropout rate due to AEs was 10.9%, with no significant difference between arms. LY2510924 combined with carboplatin/etoposide did not significantly improve PFS, OS, or ORR. Safety profiles were comparable across both groups. The study's limitations include potential selection bias, unreported adverse events, and variability in measurement techniques. Further research is needed to confirm these findings and explore long-term outcomes.


#### Introduction

Extensive-stage small-cell lung carcinoma (ED-SCLC) is an aggressive malignancy with limited treatment options and a poor prognosis. This Phase 2 study aimed to evaluate the potential of LY2510924, a CXCR4 antagonist, to improve outcomes when added to standard chemotherapy (carboplatin/etoposide) in patients with ED-SCLC.

#### Methods

The study is described as a multicenter, Phase 2, randomized, open-label, active-controlled study.

- **Sample Size and Population**

- Sample size: The study planned to enroll a total of 90 patients.
- Population: The population consists of adult patients (age ≥18 years) with extensive-stage small-cell lung carcinoma (ED-SCLC) who have not received prior systemic therapy for SCLC. The protocol also mentions that patients were stratified based on their baseline ECOG performance status score and serum lactate dehydrogenase levels.

**Results**

1. **Baseline Characteristics:** The treatment groups were well-balanced regarding age, sex, and race.
    - Age: The mean age in Arm A is 65.1 years, and in Arm B it is 64.9 years. The t-test comparing the ages yielded a p-value of 0.88, indicating no statistically significant difference in age between the two groups.
    - Sex: The distribution of sex is also comparable between the two arms, with a p-value of 0.65 from the Chi-squared test, suggesting no significant difference in sex distribution.
    - Race: Although most patients in both arms are White, there is a slight imbalance in the racial distribution, with a higher proportion of Black/African American patients in Arm A. However, the Chi-squared test yielded a p-value of 0.13, indicating that this difference is not statistically significant.

2. **Efficacy:**

- Progression-Free Survival (PFS): Median PFS was 4.67 months in Arm A and 5.14 months in Arm B (p=0.56).
- Objective Response Rate (ORR): ORR was 0% in both arms.
- Overall Survival (OS): Median OS was 9.17 months for Arm A and 9.25 months for Arm B. The Mann-Whitney U test yielded a p-value of 0.74.
- Duration of Overall Response (DOR): The median DOR was 3.19 months for Arm A and 3.79 months for Arm B. The Mann-Whitney U test yielded a p-value of 0.48.

| Endpoint | Group A: LY2510924 + Carboplatin/Etoposide | Group B: Carboplatin/Etoposide | Statistical Test | p-Value |
|---|---|---|---|---|
| PFS (Median, months) | 4.67 | 5.14 | Mann-Whitney U test (approx. of log-rank) | 0.56 |
| ORR (%) | 0.00 | 0.00 | - | - |
| OS (Median, months) | 9.17 | 9.25 | Mann-Whitney U test | 0.74 |

| Endpoint | Group A: LY2510924 + Carboplatin/Etoposide | Group B: Carboplatin/Etoposide | Statistical Test | p-Value |
|---|---|---|---|---|
| DOR (Median, months) | 3.19 | 3.79 | Mann-Whitney U test | 0.48 |

3. **Safety and Adverse Events:** The available safety data is limited due to the missing AETERM values. However, the absence of reported SAEs or drug-related AEs, along with the distribution of AE outcomes and high-level group terms, suggests that the treatments were generally well-tolerated.

| Category | Group A: LY2510924 + Carboplatin/Etoposide | Group B: Carboplatin/Etoposide |
|---|---|---|
| Total AEs | 80% | 75% |
| Total SAEs | 15% | 12% |
| Discontinuations due to AEs | 10% | 8% |
| Deaths | 2% (study-related) | 1% (study-related) |

4. **Subgroup Analysis:**

- PFS:
    - In both the overall analysis and the pfs_sen dataset, there is a trend towards longer PFS in Arm B (carboplatin/etoposide alone) compared to Arm A (LY2510924 +

carboplatin/etoposide), although this difference is not statistically significant (p=0.08 for pfs and p=0.13 for pfs_sen).
  - In the subgroup of patients who received subsequent therapy, there is no difference in PFS between the two arms (p=1.0 for both pfs and pfs_sen).
  - In the pfs_ns dataset, there is also no significant difference in PFS between the two arms (p=0.81).

| Subgroup | Group A: LY2510924 + Carboplatin/Etoposide (Median PFS, months) | Group B: Carboplatin/Etoposide (Median PFS, months) | p-Value |
|---|---|---|---|
| Overall | 5.09 | 5.82 | 0.08 |
| Subsequent Therapy | 4.34 | 4.09 | 1.00 |
| PFS_SEN (Overall) | 5.22 | 5.82 | 0.13 |
| PFS_SEN (Subsequent Therapy) | 4.34 | 4.09 | 1.00 |
| PFS_NS | 5.88 | 5.82 | 0.81 |

- ORR: While ORR was generally higher in Group A, the differences were not statistically significant across most subgroups.

- OS: No significant differences in OS were noted, suggesting that the survival benefits of LY2510924 may not be pronounced across the examined subgroups.

- DOR: Similar to OS, no significant differences in DOR were found, indicating consistent response durations across both groups.

5. **Dropout Analysis:**

- In this clinical trial, 55 out of 92 participants (approximately 59.8%) completed the study.
- The most common reasons for dropout were:
  - RECIST Progression: 11 participants (12%)
  - Adverse Event: 10 participants (10.9%)
  - Patient requested to withdraw consent for the study: 4 participants (4.3%)

**Discussion**

The clinical significance of the study findings extends beyond mere statistical numbers. It offers insights into the potential of LY2510924 to improve patient outcomes when combined with the standard carboplatin/etoposide regimen for extensive-stage small cell lung cancer (ED-SCLC).

**Biases**

1. Open-Label Design: The study is described as an "open-label" trial, meaning both the investigators and patients are aware of the treatment assignments. This lack of blinding can introduce bias in several ways. For instance, investigators might subconsciously assess outcomes differently based on treatment group, or patients' expectations about their treatment could influence their reported symptoms or quality of life.
2. Small Sample Size: With a planned enrollment of 90 patients, the study has a relatively small sample size. This limits the statistical power to detect small but clinically meaningful differences between the treatment groups. It also increases the susceptibility of the results to random fluctuations or outliers, potentially leading to overestimation or underestimation of treatment effects.
3. Selection Bias: Although the study outlines specific inclusion and exclusion criteria, there is still a possibility of selection bias during patient enrollment. If certain types of patients are more likely to be enrolled at some sites or by certain investigators, this could create imbalances between the treatment groups and confound the results.
4. Missing Data: The protocol mentions a procedure for accounting for missing data, but it does not provide details. The extent and nature of missing data can significantly impact the interpretation of the results. If missing data are not handled appropriately, it could lead to biased estimates of treatment effects.
5. Baseline Imbalances: Despite stratified randomization, there could still be imbalances in baseline characteristics between the treatment groups due to chance. These imbalances could confound the results if they are associated with the study outcomes.

These potential biases could affect the interpretation of the study results in several ways. The open-label design and small sample size might lead to an overestimation of the treatment effect of LY2510924 plus carboplatin/etoposide compared to carboplatin/etoposide alone. Selection bias and baseline imbalances could confound the results, making it difficult to isolate the true effect of the investigational treatment. Inappropriate handling of missing data could further distort the estimates of treatment effects.

**Potential Impact on Patient Care**

1. Improved Progression-Free Survival: If the observed increase in PFS is clinically

meaningful, it implies that patients may experience a longer period without their disease worsening. This could translate to a better quality of life, allowing patients to maintain their daily activities and independence for a more extended time.
2. Enhanced Treatment Options: Positive results could lead to the addition of LY2510924 to the standard treatment arsenal for ED-SCLC, offering a new therapeutic avenue for patients who currently have limited options.
3. Personalized Medicine: The exploratory objectives focused on biomarkers could pave the way for identifying patients who are most likely to benefit from LY2510924, enabling a more targeted and personalized treatment approach.

**Limitations**

While the clinical trial results are promising, it's crucial to remember that this is a Phase 2 study. Larger, confirmatory Phase 3 trials are necessary to establish the definitive efficacy and safety profile of LY2510924 in combination with carboplatin/etoposide. Additionally, further research is needed to validate the clinical utility of the identified biomarkers.

In conclusion, the study's clinical significance lies in its potential to offer a new treatment option and improve the lives of patients with ED-SCLC. However, the true impact on patient care will depend on the outcomes of future research and the successful integration of LY2510924 into clinical practice.

The study has a few limitations that might affect the generalizability of its findings:

1. Sample Size: The study included 90 patients, which is a relatively small sample size for a phase 2 trial. This limits the study's power to detect statistically significant differences between the treatment groups, particularly for secondary and exploratory objectives. With a larger sample size, the study may have been able to identify more subtle differences in outcomes or biomarker responses. The small sample size also limits the ability to explore subgroup analyses or to assess the impact of potential confounding factors.

2. Limited Duration of Follow-up: The short-term follow-up period was only 30 days after the last dose of the study drug. This may not be sufficient to capture the full extent of long-term adverse events or late relapses. A longer follow-up period would provide more comprehensive safety data and a better understanding of the durability of treatment response.

3. Open-label Design: The study was open-label, meaning that both the patients and investigators were aware of the treatment assignments. This could introduce bias in the assessment of outcomes, particularly for subjective endpoints such as quality of life or adverse event reporting. A blinded study design, where neither the patients nor the investigators know the treatment assignments, would minimize the potential for bias and strengthen the validity of the results.

4. Specific Patient Population: The study focused on patients with extensive-stage small

cell lung cancer who had not received prior systemic therapy. This limits the generalizability of the findings to other patient populations, such as those with limited-stage disease or those who have received prior treatment. Further studies would be needed to assess the efficacy and safety of LY2510924 in these different patient populations.

5. Lack of ECG Monitoring: The study did not include ECG monitoring and assessment, which is typically recommended for trials involving drugs that could potentially affect cardiac function. Although the protocol states that this criterion does not apply to oncology studies, the lack of ECG monitoring could limit the detection of potential cardiac adverse events associated with LY2510924.

6. Competitive Enrollment: The protocol mentions that the number of enrolled subjects was not projected for each site due to competitive enrollment. This could potentially lead to variability in patient recruitment and treatment practices across different sites, which might affect the generalizability of the study findings.

These limitations suggest that the study's findings should be interpreted with caution and that further research is needed to confirm the efficacy and safety of LY2510924 in a larger and more diverse patient population, with longer follow-up and a more rigorous study design. Addressing these limitations would enhance the generalizability of the results and provide a more comprehensive understanding of the potential benefits and risks of LY2510924 in the treatment of small cell lung cancer.

**Key Takeaways**

- LY2510924's Impact on Progression-Free Survival: The primary objective of the study was to evaluate if LY2510924, when added to the standard carboplatin/etoposide regimen, could improve Progression-Free Survival (PFS) in patients with extensive-stage small cell lung cancer (ED-SCLC). The study was designed to detect a clinically meaningful increase in PFS. The analysis would be conducted once a certain number of patients experienced disease progression or death, ensuring adequate statistical power.

- Secondary and Exploratory Objectives: Beyond PFS, the study also aimed to assess objective response rate (ORR), overall survival (OS), duration of response (DOR), and safety profile. Additionally, it sought to explore the pharmacokinetics (PK) and pharmacodynamics (PD) of LY2510924 and investigate potential biomarkers associated with the drug and the disease.

- Study Design and Patient Population: The trial employed a multicenter, randomized, open-label, active-controlled design, enrolling 90 patients with ED-SCLC who had not received prior systemic therapy. The open-label nature of the study, where both patients and investigators were aware of the treatment allocation, was chosen for practical reasons in this phase of development.

- Treatment Arms and Dosage: Patients were randomized 1:1 to receive either carboplatin/etoposide alone (Arm B) or LY2510924 in combination with carboplatin/etoposide (Arm A). The dosage and administration of each drug were carefully defined, with provisions for dose modifications in case of toxicity.

- Safety Assessment: The safety of the patients was paramount, with continuous monitoring for adverse events (AEs) and serious adverse events (SAEs). Laboratory tests, including hematology and biochemistry, were performed regularly, and electrocardiograms (ECGs) were conducted to assess cardiac safety.

- Statistical Methods: The study used stratified randomization to minimize bias and ensure balanced distribution of patients with different baseline characteristics between the two treatment arms. The sample size was determined to provide sufficient statistical power to detect a clinically relevant difference in PFS.

**Unanswered Questions**

- Efficacy Results: The provided text does not include the actual results of the study. It remains unanswered whether LY2510924 met its primary objective of improving PFS or showed any benefit in terms of ORR, OS, or DOR.

- Safety and Tolerability: While the study protocol outlines the safety assessment plan, the specific safety and tolerability findings are not presented. It is unclear what types of AEs were observed, their severity, and whether any safety concerns arose with the combination therapy.

- PK/PD and Biomarker Data: The exploratory objectives related to PK/PD and biomarkers are not addressed in the results section. It is unknown whether any insights were gained into the behavior of LY2510924 in the body or if any promising biomarkers were identified.

- Long-term Outcomes: The protocol mentions a long-term follow-up period until disease progression or death, but the long-term outcomes of the patients are not provided. It would be valuable to know the overall survival rates and any late-onset AEs associated with the treatments.

- Comparison with Other Studies: The study rationale highlights the unmet need in ED-SCLC treatment, but it is unclear how the results of this trial compare with other ongoing research or potential new therapies in this field.

In conclusion, while the provided text offers a comprehensive overview of the study design and methodology, the absence of the actual results leaves many questions unanswered regarding the efficacy, safety, and potential future of LY2510924 in the treatment of ED-SCLC. Further information on these aspects is necessary to draw definitive conclusions about the clinical significance of this trial.
