## Supplementary 5 - Investigator4 Report for "Artificial Intelligence in Biomedical Data Analysis: A Comparative Assessment of Large Language Models for Automated Clinical Trial Interpretation and Statistical Evaluation"

**Report: Analysis of LY2510924 Combined with Carboplatin/Etoposide Versus Carboplatin/Etoposide in SCLC**

**1. Introduction**

This analysis evaluates the efficacy and safety of LY2510924 combined with carboplatin/etoposide (Arm A) versus carboplatin/etoposide alone (Arm B) in patients with extensive-stage small cell lung cancer (SCLC). Data from various endpoints—progression-free survival (PFS), overall survival (OS), objective response rate (ORR), and adverse events (AEs)—were analyzed.

**2. Baseline Characteristics**

A review of the baseline characteristics indicated that both treatment arms were largely comparable in terms of age, sex distribution, and other baseline health markers. However, ECOG performance status data was missing, which limited a full assessment of the comparability of patients' functional status.

**3. Progression-Free Survival (PFS)**

- **Median PFS**:
  - **Arm A**: 4.90 months
  - **Arm B**: 5.32 months
- **Mean PFS**:
  - **Arm A**: 5.26 months
  - **Arm B**: 5.62 months
- **P-Value (Statistical Test)**: 0.2528 (not statistically significant)

The slight advantage of Arm B in terms of PFS was not statistically significant. This suggests that the combination of LY2510924 with chemotherapy does not provide a substantial improvement in delaying disease progression.

| Treatment Arm | Median PFS (months) | Mean PFS (months) | P-Value |
|---|---|---|---|
| LY2510924 + Carboplatin/Etoposide (Arm A) | 4.90 | 5.26 | 0.2528 |
| Carboplatin/Etoposide (Arm B) | 5.32 | 5.62 | |

**4. Overall Survival (OS)**

- **Median OS**:
  - **Arm A**: 9.17 months
  - **Arm B**: 9.25 months
- **Mean OS**:

- **Arm A**: 8.60 months
  - **Arm B**: 10.48 months
- **P-Value (Statistical Test)**: 0.0806 (not statistically significant)

Although Arm B showed a slightly longer overall survival, this difference was not statistically significant.

| Treatment Arm | Median OS (months) | Mean OS (months) | P-Value |
|---|---|---|---|
| LY2510924 + Carboplatin/Etoposide (Arm A) | 9.17 | 8.60 | 0.0806 |
| Carboplatin/Etoposide (Arm B) | 9.25 | 10.48 | |

**5. Objective Response Rate (ORR)**

- **ORR**:
  - **Arm A**: 83.87%
  - **Arm B**: 90.41%
- **P-Value (Chi-Square Test)**: 0.1293 (not statistically significant)

While Arm B had a higher ORR, this difference was not statistically significant, indicating comparable tumor response rates between the two treatment arms.

| Treatment Arm | ORR (%) | P-Value |
|---|---|---|
| LY2510924 + Carboplatin/Etoposide (Arm A) | 83.87 | 0.1293 |
| Carboplatin/Etoposide (Arm B) | 90.41 | |

**6. Adverse Events (AEs)**

The most common adverse events were similar between the two arms, including anemia, fatigue, and nausea. Both arms showed high rates of side effects, but the addition of LY2510924 did not substantially increase toxicity.

| Adverse Event | Arm A (%) | Arm B (%) |
|---|---|---|
| Anemia | 206.38 | 193.18 |
| Fatigue | 119.15 | 138.64 |
| Thrombocytopenia | 123.40 | 120.45 |
| Nausea | 110.64 | 104.55 |
| Neutropenia | 93.62 | 113.64 |

**7. Safety Concerns**

Both treatment arms demonstrated high rates of common chemotherapy-related toxicities, such as anemia, fatigue, and neutropenia. Importantly, the safety profiles of the two treatment arms were comparable, with no major safety concerns arising from the addition of LY2510924.

**8. Conclusion**

- **Efficacy**: The addition of LY2510924 to standard chemotherapy did not show statistically significant improvements in PFS, OS, or ORR. The slight differences favoring Arm B (carboplatin/etoposide) were not large enough to suggest a substantial benefit from adding LY2510924.
- **Safety**: Both treatment arms had similar adverse event profiles, with common side effects such as anemia, fatigue, and nausea. No unexpected safety concerns were identified with LY2510924.
- **Clinical Implications**: Given the lack of significant efficacy advantages and the comparable safety profile, the addition of LY2510924 to standard chemotherapy may not provide meaningful clinical benefit for most patients with extensive-stage SCLC. The standard carboplatin/etoposide regimen remains a viable option.

**9. Unanswered Questions**

- The missing ECOG performance status data limits the ability to fully assess whether the two groups were comparable in terms of baseline functional status.
- Further research is needed to explore potential long-term survival benefits, as well as to identify specific patient subgroups who may respond more favorably to LY2510924.
