## Supplementary Information 1 for "Artificial Intelligence in Biomedical Data Analysis: A Comparative Assessment of Large Language Models for Automated Clinical Trial Interpretation and Statistical Evaluation"

#### 1.0 METHODS

The CoT prompting provided a structured framework that guided the LLMs through a systematic evaluation of the clinical trial data. The template included the following key steps:

**Define the Study Objective**: The LLMs were prompted to identify the primary research question and secondary outcomes of interest, focusing on the efficacy of LY2510924 in combination with standard chemotherapy agents.

Key questions for defining the study objective:

a) What's the primary research question?
b) What are the secondary outcomes of interest?

**Review Study Design**: The models assessed the randomized, controlled nature of the study, including sample size and population characteristics, to understand the experimental setup.

Key questions for reviewing the study design:

a) Is it randomized? Blinded? Controlled?
b) What's the sample size and population characteristics?

**Examine Data Quality**: LLMs evaluated the dataset for missing data points and how outliers were handled, ensuring the integrity and reliability of the data.

Key questions for examining data quality:

a) Are there missing data points?
b) How were outliers handled?

**Analyze Baseline Characteristics**: The models compared treatment groups to determine if they were comparable at baseline and identified any significant differences that could influence outcomes.

Key questions from analyzing baseline characteristics:

a) Are treatment groups comparable?
b) Any significant differences that could impact results?

**Evaluate Primary Outcome**: LLMs calculated the main effect size for PFS, assessed statistical significance, and determined confidence intervals to evaluate efficacy.

Key questions from evaluating primary outcome:

a) What's the main effect size?
b) Is it statistically significant?
c) What's the confidence interval?

**Assess Secondary Outcomes**: The models analyzed secondary endpoints like OS, ORR, and DOR to see if they supported the primary findings and noted any unexpected results.

Key questions assessing secondary outcomes:

a) Do they support the primary outcome?
b) Any unexpected findings?

**Consider Subgroup Analyses**: LLMs explored effects across different subgroups (e.g., age, gender, disease stage) to identify any consistent patterns or notable differences.

Key questions while considering subgroup analyses:

a) Are effects consistent across subgroups?
b) Any notable differences?

**Examine Safety Data**: The models reviewed adverse events and serious safety concerns to assess the treatment's risk profile.

Key questions while examining safety data:

a) What adverse events occurred?
b) Any serious safety concerns?

**Analyze Dropout Rates**: LLMs assessed participant retention, noting how many completed the study and reasons for any dropouts, which could affect the study's validity.

Key questions while analyzing dropout rates:

a) How many participants completed the study?
b) Reasons for dropouts?

**Consider Potential Biases**: The models identified factors that could skew results, such as selection bias or confounding variables, and discussed their impact on interpretation.

Key questions while considering potential biases:

a) Any factors that could skew results?
b) How might these impact interpretation?

**Evaluate Statistical Methods**: LLMs reviewed the appropriateness of the statistical tests used and how multiple comparisons were managed to ensure robust conclusions.

Key questions evaluating statistical methods:

a) Were appropriate tests used?
b) How were multiple comparisons handled?

**Compare to Similar Studies**: The models compared findings with existing literature to contextualize results within the broader field of SCLC treatment research.

Key questions comparing to similar studies:

a) How do results align with previous research?
b) Any contradictions or new insights?

**Assess Clinical Significance**: Beyond statistical significance, LLMs considered the practical impact of the findings on patient care and treatment protocols.

Key questions assessing clinical significance:

a) Beyond statistical significance, what's the practical impact?
b) How might this affect patient care?

**Consider Limitations**: The models identified weaknesses in the study, such as sample size or data limitations, discussing how these might affect generalizability.

Key questions considering limitations:
a) What are the study's weaknesses?
b) How might these affect generalizability?

**Formulate Conclusions**: Finally, LLMs synthesized the analysis to highlight key takeaways and identified areas for future research.

Key questions for formulateing conclusions:
a) What are the key takeaways?
b) What questions remain unanswered?

To ensure consistency and reduce variability in the analysis of the chosen clinical trial, we standardized the analytical approach across different Large Language Models (LLMs) and investigators. This was achieved by templating the workflow of clinical trial reporting and tuning the prompts to generate a standardized report.

### 2.0) RESULTS

**Survival Curves for PFS and OS (Generated by Code Generated by LLM)**

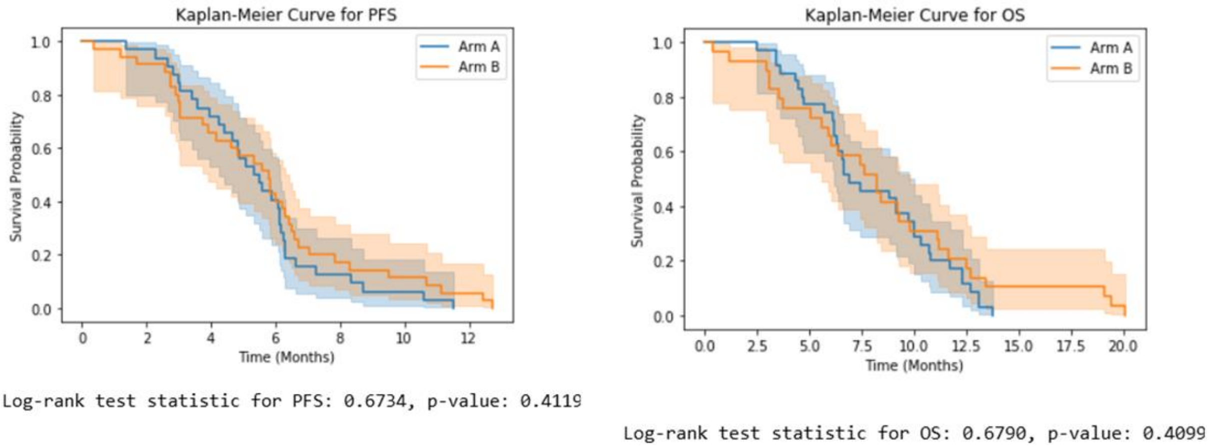


**Figure S1.** Kaplan-Meier survival curves illustrating Progression Free Survival (PFS) and Overall Survival (OS). Curves were generated using code produced by an LLM based on data from a clinical trial of patients with ES-SCLC treated with LY2510924 plus carboplatin/etoposide (Arm A) or carboplatin/etoposide alone (Arm B).
